# Epigenetic, ribosomal, and immune dysregulation in Paediatric Acute-Onset Neuropsychiatric Syndrome

**DOI:** 10.1101/2025.03.28.25324649

**Authors:** Velda X Han, Sarah Alshammery, Brooke A Keating, Brian S Gloss, Markus J Hofer, Mark E Graham, Nader Aryamanesh, Lee L Marshall, Songyi Yuan, Emma Maple-Brown, Jingya Yan, Sushil Bandodkar, Kavitha Kothur, Hiroya Nishida, Hannah Jones, Erica Tsang, Xianzhong Lau, Ruwani Dissanayake, Iain Perkes, Shekeeb Mohammad, Fabienne Brilot, Wendy Gold, Shrujna Patel, Russell C Dale

## Abstract

Paediatric Acute-Onset Neuropsychiatric Syndrome (PANS) is characterised by abrupt onset obsessive compulsive disorder and regression in neurodevelopmental skills, triggered by infection or stress. Whether PANS is a distinct entity or part of a neurodevelopmental spectrum is uncertain, and its pathophysiology remains unclear. We show that children with PANS (n=32) and other non-PANS (n=68) neurodevelopmental disorders (total n=100) have higher reported early childhood infections and a loss of previously acquired developmental skills compared to neurotypical controls (n=58). Children with PANS have normal routine immune testing, however bulk RNA-sequencing (PANS n=20 vs controls n=15) revealed upregulated pathways in ribosomal biogenesis and RNA methyltransferases, and downregulated pathways in diverse cellular functions such as mitochondrial activity, cell signalling, endocytosis, and immune responses. Single-cell RNA-sequencing (PANS n=2 vs controls n=2) confirmed these findings but showed heterogeneity across immune cell types. Toll-like receptor stimulation assay using peripheral blood mononuclear cells revealed reduced TNF and interleukin-6 responses in PANS patients (n=7) compared to controls (n=7). RNA sequencing before and after intravenous immunoglobulin treatment in PANS patients (n=9 vs controls n=10) revealed reversal of the dysregulated ribosomal, epigenetic, and cell signaling pathways. Given the central role of the immune system in synaptic pruning and neurodevelopment, these insights provide rationale for novel epigenetic and immune modulating therapies to optimize neurodevelopmental trajectories and minimize neuropsychiatric impairment.

## Introduction

Neurodevelopmental disorders (NDDs) including autism spectrum disorder (ASD), attention deficit hyperactivity disorder (ADHD), Tourette syndrome/tics disorder, and obsessive-compulsive disorder (OCD) are increasing in prevalence, and now affect 1 in 6 children^1^. Overlaps in clinical symptoms and comorbidities of NDDs suggest shared genetic etiologies^2^. While genetic factors exert a significant influence on NDDs, most cases are attributed to common susceptibility loci, rather than rare gene variants^2, 3^. The interplay of genetic and environmental factors in the pathogenesis of childhood NDDs is increasingly recognized, starting from preconception and extending into early adulthood^4^. During this period, the central nervous system undergoes dynamic changes including neurogenesis, cell migration, cell differentiation, and synaptogenesis. Additionally, brain immune cells, primarily microglia, actively refine neural networks, through synaptic pruning^5, 6^. Early life programming, influenced by environmental exposures during critical developmental periods, can affect a child’s developmental trajectory^4^. This lasting impact is proposed to occur through cellular stress on the epigenetic and gene regulatory program of brain and immune cells ^7, 8^.

Environmental exposures in the postnatal period, such as infections and stress are known to trigger or cause fluctuations in tics, or neuropsychiatric symptoms in children with NDDs^9–12^. However, a small subgroup of children can suffer abrupt and severe onset of neuropsychiatric symptoms accompanied by developmental regression, leading to the proposal of clinical syndromes, such as Paediatric acute-onset neuropsychiatric syndrome (PANS) or Paediatric autoimmune neuropsychiatric disorder associated with Streptococcal infection (PANDAS), which is a subset of PANS ^13, 14^. Although these syndromes have been described in literature, their aetiology remains enigmatic, thus not fully recognized as ‘discrete biological entities’. Early clinical observations of PANS/PANDAS proposed these conditions as distinct autoimmune entities, akin to autoimmune encephalitis ^14^. However, an alternative hypothesis is that PANS is part of the broader NDD continuum, representing a phenotype that demonstrates ongoing interactions between the immune system and brain in a subset of people with NDDs^15^. Studies that investigate the underlying pathophysiology of PANS/PANDAS are lacking. Furthermore, antibiotics and immune modulation, such as intravenous immunoglobulin (IVIg), have been proposed to benefit individuals with PANS/PANDAS, but these treatments remain contentious ^16, 17^.

We developed a program to explore the influence of environmental factors, in particular recurrent infections, on neuropsychiatric symptoms, as well as associated loss of developmental skills. We focussed on children who fulfil criteria for PANS, who have abrupt infection or stress triggered neuropsychiatric symptoms, as a model to examine gene-environment interactions in NDDs (Figure 1A). We hypothesize that epigenetic and immune processes operate at the gene-environment interface and are central to the pathogenesis of PANS and other NDDs. We performed bulk blood transcriptomic analyses in PANS, and then further explored these findings in PANS via single cell transcriptomics and functional immune assays. We also conducted blood transcriptomic analysis on children with PANS both before and after intravenous immunoglobulin administration, aiming to define the biological effect of IVIg in these individuals.

**Figure 1.**
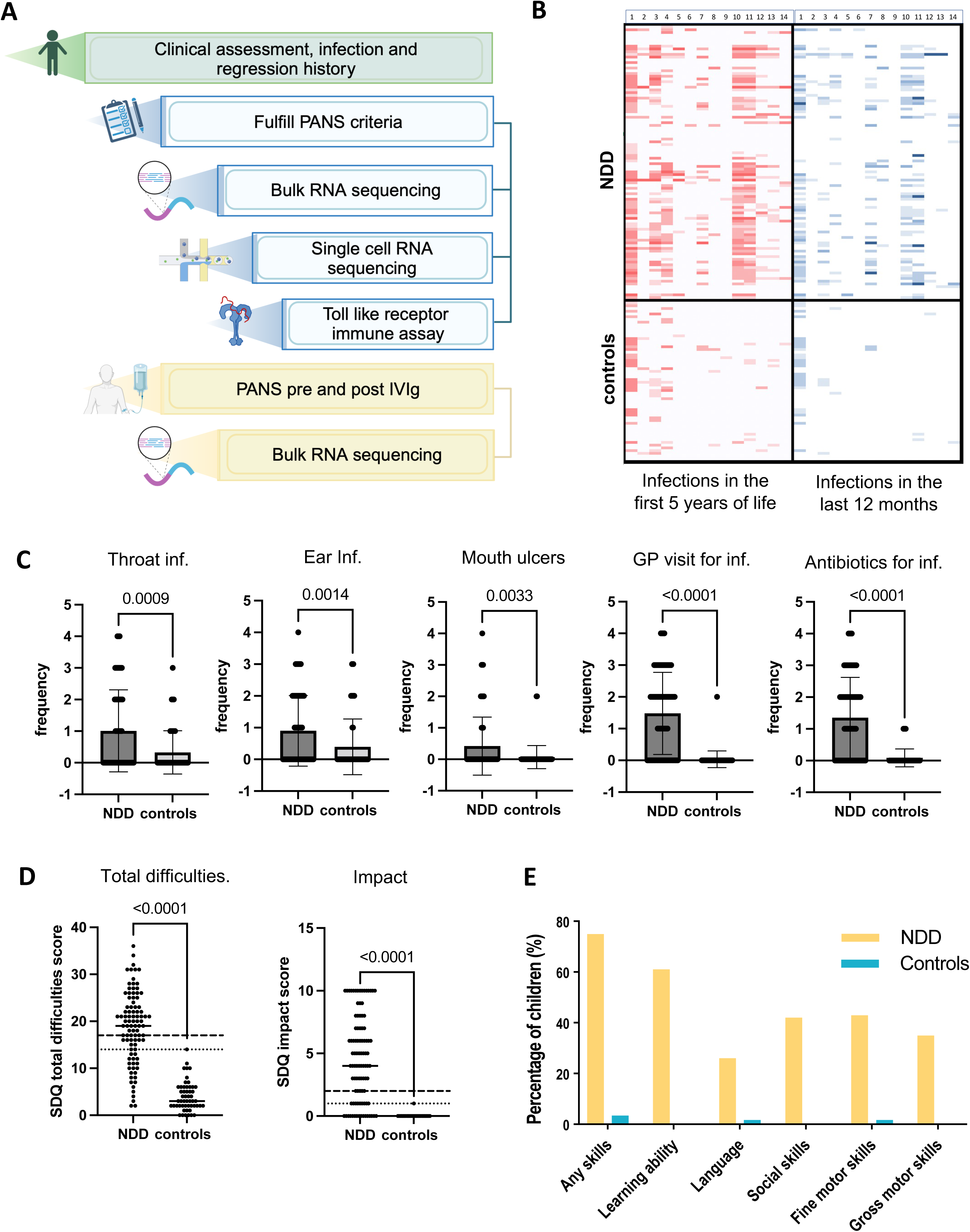
Clinical data of 100 children with neurodevelopmental disorders (NDD) versus 58 healthy controls. **(A)** The study workflow involved clinical interview of children with NDDs and controls. Bulk blood RNA sequencing, single-cell blood RNA sequencing, and Toll-like receptor stimulation assay were performed for children with Paediatric acute-onset neuropsychiatric syndrome (PANS). Children with PANS receiving intravenous immunoglobulin (IVIg) treatment were categorized as the PANS-IVIg cohort. Bulk blood RNA sequencing was performed before and after IVIg treatment. Image generated with *Biorender*. **(B)** Heatmap of reported childhood infections of 100 children with NDDs (top half) versus 58 healthy controls (bottom half). Responses to infection screening tool in the first 5 years of life (in red) and 12 months prior to interview (in blue). On the x axis, numbers 0 to 14 represent the 14 questions in the infection screening tool (Supplementary Figure 1). The intensity of the colour represents the frequency based on the 5-point Likert scale. There is increased intensity in the heatmap in the children with NDDs versus controls reflecting higher frequency of reported infections in those with NDDs, particularly in the first 5 years of life. **(C)** Bar charts of frequency of reported infection in the first 5 years of life in children with NDDs versus healthy controls. The frequency is based on a 5-point Likert scale including - never, occasional (less than once per year), sometimes (1-3 times per year), often (4-6 times per year) and almost always (almost always). In the first 5 years of life, there were significantly higher rates of throat infection, ear infection, mouth ulcers, general practitioner visits for infection, antibiotic courses and emergency visits for infection in the NDD group compared to controls. **(D)** Dot plots showing the total difficulties and impact score of the Strengths Difficulties Questionnaire (SDQ). Children with NDDs compared to healthy controls have a higher average total difficulties and impact score (p<0.0001). **(E)** Bar plot showing the percentage of children with NDDs (in yellow) and controls (in blue) with reported loss of skills. A significantly higher proportion of children with NDDs compared to healthy controls report loss of skills at any time in their childhood, including learning ability, social skills, fine and gross motor skills.

## Methods

### Participant selection

#### NDD cohort

We performed a prospective study of 100 sequential children (<18 years) referred with complex NDDs to a specialist clinic at the Children’s Hospital at Westmead, Sydney from January 2020 to January 2023. In this clinic, there is a referral enrichment of PANS, Tourette syndrome, and OCD, often with accompanying disorders, such as ASD and ADHD. Children seen at this clinic are screened using a structured clinical assessment on REDCap, followed by a clinical directed interview.

#### PANS cohort

Within the NDD cohort, 32 children met the criteria for PANS ^13^. Additionally, 4 more children who met the criteria for PANS were recruited prospectively. We used the 2013 PANS diagnostic criterion which included an abrupt, dramatic onset of OCD or severely restricted food intake^13^. There was concurrent presence of at least two of the seven categories including (i) anxiety, (ii) emotional lability and/or depression, (iii) irritability, aggression, and/or oppositional behaviours, (iv) behavioural regression, (v) deterioration in school performance, (vi) sensory or motor abnormalities, (vii) somatic signs and symptoms, including sleep disturbances, enuresis or urinary frequency^14^. Abrupt and dramatic onset of neuropsychiatric symptoms was defined as within 48 hours. Children with PANS included in the biological investigations all had new or ongoing neuropsychiatric symptoms (Supplementary Table 1) ^13^.

#### PANS-IVIg cohort

Within the PANS cohort, a subgroup of 9 children were about to commence or were receiving ongoing open-label intravenous immunoglobulin (IVIg) treatment (1.5- 2g/kg per month) ^14^. In Australia, PANS is currently a Medicare-approved indication for IVIg.

These patients who received IVIg had debilitating neuropsychiatric symptoms, despite optimization of psychology and conventional psychiatric treatments.

#### Controls

We recruited age- and sex- matched healthy children of hospital workers or patients being investigated for non-neurological and non-inflammatory disorders (eg. short stature) (Supplementary Table 2). We screened 58 healthy controls using the same structured clinical assessment on REDCap as the NDD patients. Within this group, 29 controls were involved in biological investigations. The inclusion criterion for controls involved in biological investigations was the absence of NDDs, autoimmune diseases and severe allergic conditions. Four controls had previous neurological symptoms (epilepsy in 3, benign intracranial hypertension in 1) that were resolved prior to blood taking.

All children with PANS and controls used in these biological blood studies were screened to ensure absence of any infections, allergic reactions/anaphylaxis, other major medical conditions, or admissions in the four weeks before blood draw.

### Structured clinical assessment on REDCap and clinical directed interview

A structured assessment using REDCap was completed by parents of participants. Childhood infection history was captured using a purpose-built screening tool for infection (Supplementary Figure 1). There is currently no standardised tool to quantify infection rates in children, and most questionnaires are designed to screen for immunodeficiencies. Thus, we developed a screening tool that records frequencies of common viral and bacterial infections in childhood, involving respiratory tract, urinary tract, recurrent mouth ulcers, skin infections, and more serious bacterial infections (e.g. pneumonia, meningitis, bone infections). Additional questions on frequency of visits to general practitioners, emergency department, or hospitalisations due to infection, and frequency of antibiotic courses, were also included. A 5-point Likert scale was used to assess frequency of these infections, with options including never, occasional (less than once per year), sometimes (1-3 times per year), often (4-6 times per year) and almost always (so frequent, hard to count), in the first 5 years of life and the last 12 months of life (scored 0-4). Objective scores of functioning, including the Strengths and Difficulties Questionnaire (SDQ), was performed for each child ^18^. The presence or absence of loss of skills at any time were also recorded (eg. loss of learning ability attention/concentration, language, social skills, fine and gross motor skills).

For the NDD cohort, the clinicians (RCD, SSM, VXH) performed a clinical directed interview and recorded the child’s diagnosis using DSM-5 criteria, including PANS criteria^13, 19^. In addition, markers of disease severity, including emergency department attendance due to neurodevelopmental symptoms, and absence from school for more than three months, were recorded. Triggers (e.g., stress, infection, excitement) that worsen neurodevelopmental symptoms were also recorded.

For the PANS-IVIg cohort, we used the Clinical Global Impression Scale-Severity score (CGI-S) as a quantitative measure for the therapeutic response to IVIg in PANS, and charted their daily CGI-S scores over four-week cycles^20^. The CGI-S question is: “Considering your total clinical experience with this particular population, how mentally ill is the patient at this time?”^20^. This rating is based upon observed and reported symptoms, behaviour, and function. A seven-point rating scale is used which ranged from 1=normal to 7= extremely ill.

### Sample collection

After written consent, venous blood samples were collected for testing in PAXgene™ blood RNA tubes (Qiagen, Hilden, Germany) and Acid Citric Dextrose tubes (BD).

### Bulk blood RNA-sequencing

We performed bulk blood RNA sequencing in the PANS cohort comprising 20 children with PANS versus 15 gender and age-matched controls. There were no significant differences in age, gender or clinical characteristics between the 20 children with PANS who underwent bulk blood RNA sequencing, versus the other 12 in the cohort. Subsequently, we performed bulk blood RNA sequencing in the PANS-IVIg cohort comprising another 9 children with PANS before and after IVIg treatment versus 10 gender and age-matched controls. Blood samples were collected immediately prior to IVIg administration, and 10-14 days post IVIg. In the PANS-IVIg cohort, bulk RNA sequencing was performed via two batches, 4 patients versus 4 controls in the first batch, and 5 patients versus 6 controls in the second batch. There was no addition of new medications during the period when omics analyses were performed. Bulk blood RNA sequencing was conducted by the Australian Genome Research Facility (AGRF, Melbourne, Australia), as described in Supplementary material.

### Single-cell blood RNA sequencing

We performed single-cell RNA sequencing using the 10X Genomics platform on an additional two children with PANS who had not previously undergone bulk RNA sequencing. compared with two matched control subjects, as described in Supplementary material.

### Toll-like receptor immune assay

We developed an in-house Toll-like receptor (TLR) immune assays on PBMCs from 7 children with PANS versus 7 gender and age-matched controls, as described in Supplementary material.

## Bioinformatic analysis

### Bulk/single-cell RNA sequencing bioinformatic analysis and enrichment analysis

RNA seq data were analyzed (by VXH, SA, EM, BG, NA, ML) in the R statistical environment ^21^ with *tidyverse*^22^, described in Supplementary material. For 10X Genomics single cell RNA sequencing, the *Seurat* package was used for analysis^23^, described in Supplementary material. In this study, we focused on a pathway driven analysis, demonstrating cumulative effects of the differential expression of multiple genes, instead of focusing on individual genes. Pathway enrichment analysis was performed via Gene Set Enrichment Analysis (GSEA) to obtain enriched Gene Ontology (GO) and reactome pathways using the *fgsea* package, described in Supplementary material.

## Statistical analysis

We performed statistical analyses using GraphPad Prism v8.2.0. We compared characteristics of NDDs versus controls using independent t-tests, chi-squared tests, and Mann-Whitney U tests.

## Ethics approval

Ethical approval was granted by the Sydney Children’s Hospitals Network Human Research Ethics Committee (HREC/18/SCHN/227, 2021/ETH00356). Informed consent was obtained from all subjects.

## Results

### Neurodevelopmental disorders (NDD) cohort: clinical characteristics

A total of 100 sequentially referred children with NDDs (mean age 10.1 years 4-17), 68% males) and 58 neurotypical controls (mean age 9.5 (5-14 years), 60% males) were interviewed using the standardized assessment tool. The children with PANS phenotype were invited to contribute to biological ‘omics’ studies, after informed consent.

A low socioeconomic background was disproportionately higher in the NDD group compared to controls (median decile 8 versus 10, p<0.00001, Supplementary Figure 2). The NDD group also had significantly higher rates of gastroesophageal reflux (27% vs 1.7%, p = 0.000061), constipation (33% vs 10.3%, p=0.001), asthma (24% vs 5.2%, p=0.002), eczema (16% vs 5.2%, p=0.044), hay fever (13% vs 3.4%, p=0.048), and allergies (38% vs 5.2%, p<0.00001), compared to controls.

Parents reported significantly more frequent childhood infections in the NDD group compared to controls in first 5 years of life (in red) and last 12 months of life (in blue) (heat map presented in Figure 1B). In the first 5 years of life, there were higher rates of throat infection (p=0.0009), ear infection (p=0.0014), mouth ulcers (p=0.0033), general practitioner visits (p<0.0001), and antibiotic courses (p<0.0001) in NDDs versus controls (Figure 1C). In the 12 months prior to interview, there were higher rates of mouth ulcers (p=0.0006) and antibiotic courses (p<0.0001) in NDDs versus controls (Supplementary Figure 3).

The main diagnoses in the NDD group included tic disorder/Tourette syndrome (68%), OCD (48%), ADHD (46%), ASD (35%), plus anxiety (63%), depression (9%) and learning disability (6%). As expected, using the strengths and difficulties questionnaire (SDQ), children with NDDs had more total difficulties compared to healthy controls, and higher emotional symptoms, conduct problems, hyperactivity/inattention, and problems with peer relationships (all p<0.0001, Figure 1D and Supplementary Figure 4).

Reported environmental triggers at onset of tics/OCD were common, and occurred in 45% of the children with NDDs, including infection (26%), stress (11%) and medications (3%). Loss of skills associated with neurodevelopmental symptoms were reported in 82% of the NDD group, compared to 3.4% of controls (p<0.00001). Loss of skills in the NDD group were in the domains of learning ability (69%), fine motor skills (46%), social skills (45%), gross motor skills (37%) and language (27%) (Figure 1E). The NDD cohort were significantly impaired: 67% of children were taking one or more conventional psychiatric medication, 31% presented to emergency department due to their neuropsychiatric symptoms, and 24% had prolonged school absence (>3 months). Treatment included alpha agonist (35%), selective serotonin reuptake inhibitors (31%), neuroleptics (27%), and stimulants (27%). Immune modulators initiated or used during the time of the study included intravenous immunoglobulin (17%), antibiotics (21%), and oral corticosteroids (14%).

### PANS cohort: Clinical characteristics

Out of the 100 patients with NDDs, 32 fulfilled the PANS criteria (mean age 10.4 years (4-12) at time of study recruitment, 62.5% males)^13^. Children with PANS had predominant OCD (81.2%), restrictive eating (46.7%), and separation anxiety (84%). Additional symptoms included emotional lability/depression (68.8%), irritability/aggression/oppositional behaviours (53.1%), deterioration in school performance (87.5%), sensory or motor abnormalities (53.1%), tics (46.9%), somatic symptoms including sleep disturbance and enuresis (59.4%). A significant proportion of children with PANS had pre-existing ASD (38%), ADHD (28%), and learning disabilities (28%), prior to onset of first PANS episode (patient details in Supplementary Table 3). Although all children in the PANS cohort had a convincing history of acute onset of neuropsychiatric symptoms, infection was associated with onset in 66%, and in the other 34%, the onset was associated with manifest stress or unclear reasons. The PANS cohort was significantly impaired: 63% had prolonged school absence (>3 months), 81% report a relapsing remitting clinical course, and the mean number of conventional psychiatric drugs trialed was 2.4 (0-9). A large proportion of them previously tried infection/immune therapies including antibiotics (70%), steroids (47%) and IVIg (57%). The mean duration of time from first PANS episode to study recruitment, clinical interview and research blood sampling was 3.7 years (0.5-10 years). Although some of the children were in the chronic phase of PANS, all of them had ongoing developmental and neuropsychiatric symptoms at time of blood sampling.

### PANS versus non-PANS NDD cohort: Clinical characteristics

Children with PANS (n=32) compared to non-PANS NDDs (n=68, mean age: 10.6 years (4-18), 70.6% males) were more likely to have OCD (81% vs 37%, p=0.0001), anxiety (84% vs 57%, p=0.001), depression (69% vs 7%, p=0.00001), and more severe NDD symptoms resulting in prolonged school absence (63% vs 18%, p=0.0001) (Supplementary Table 3). There were no significant differences between childhood infection rates or loss of skills between the two groups (Supplementary Figure 3 and 4). Infections and stress triggering onset of NDD symptoms were higher in PANS compared to non-PANS NDDs (100% vs 19%, p < 0.0001). Although infection and stress were observed in non-PANS NDDs, the onset in these patients was not abrupt and dramatic, as is seen in PANS. Instead, the onset was insidious and the fluctuations ‘waxed and waned’ as is seen in Tourette syndrome, OCD, and other NDDs.

### Cerebrospinal fluid and blood inflammatory markers in PANS

Due to the infection provoked nature and abrupt onset of neuropsychiatric symptoms in PANS, the children often presented to hospital acutely, and some required cerebrospinal fluid (CSF) examination to exclude encephalitis. Within the PANS cohort, six children (mean age: 7 years (4-11), 67% males) had CSF evaluation during the first or subsequent PANS relapse. These children had negative antibody testing (NMAR, LGI1, CASPR2, GABA-BR, AMAP-R) in serum and cerebrospinal fluid. We compared CSF cytokines and inflammatory metabolites in these six children with PANS with two other control groups: (1) children with non-inflammatory neurogenetic disorders (n= 11, mean age: 6.3 years (0.3-16), 36.4%), and (2) children with proven autoimmune encephalitis (n= 8, mean age: 9 years (3-14), 62.5% males) (Supplementary Table 4). We measured CSF tumor necrosis factor (TNF), interferon (IFN)- alpha, interleukin (IL)-10 and interleukin-6 as these have been shown to be the most differentiating cytokines of encephalitis compared to controls^24^. In addition, we measured CSF neopterin and kynurenine-tryptophan ratio which are sensitive markers of neuroinflammation (described previously)^25, 26^. These CSF biomarkers were elevated in encephalitis but not elevated in PANS, and the findings in PANS did not differ from the neurogenetic ‘negative’ controls (Figure 2A). In addition, there were no quantitative cell count differences in the peripheral blood white cell, neutrophil, lymphocyte, C-reactive protein, and erythrocyte sedimentation rate in children with PANS compared to the controls (taken at time of blood omics samples, Supplementary Figure 5).

**Figure 2.**
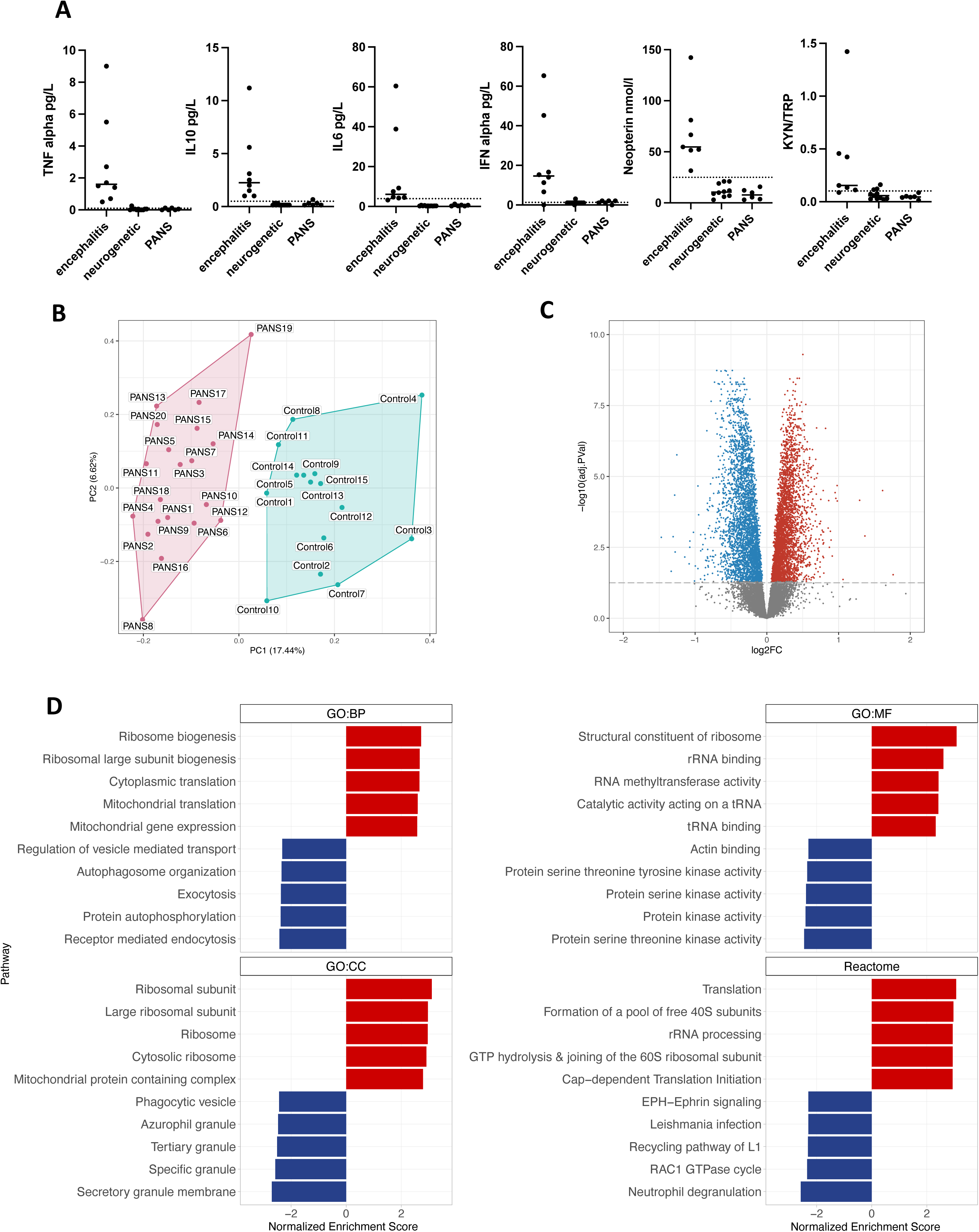
Bulk blood RNA sequencing of Paediatric acute-onset neuropsychiatric syndrome patients (PANS) versus controls. **(A)** Cerebrospinal fluid cytokine levels (TNF, IL10, IL6, IFN alpha), neopterin, and kynurenine/tryptophan (KYN/TRP) levels in children with PANS (n=6) did not significantly differ from controls with non-inflammatory neurogenetic conditions (n=11). On the other hand, children with anti-NMDA receptor encephalitis (n=8) had markedly elevated cytokine levels, neopterin and KYN/TRP compared to controls with neurogenetic conditions and PANS. The CSF findings in PANS were not in keeping with autoimmune encephalitis. **(B)** Principal component analysis (PCA) performed on bulk RNA sequencing performed in children with PANS and controls. The x-axis represents Principal Component 1 (PC1), while the y-axis represents Principal Component 2 (PC2). Unbiased hierarchical clustering of gene expression between children with PANS and controls showed separation of data indicating strong group discrimination post RUV. **(C)** Volcano plot with annotation of 6914 differentially expressed genes (3544 upregulated genes in red and 3369 downregulated genes in blue) (FDR<0.05). **(D)** Bar plot of Gene Set Enrichment Analysis (GSEA) Gene Ontology (GO) Biological Process (BP), Molecular Function (MF), Cellular Component (CC) and Reactome pathways. The top 5 upregulated GSEA GO and Reactome pathways were predominantly ribosomal biogenesis, translational processes, as well as RNA methyltransferase pathways. The top 5 downregulated GSEA GO and Reactome pathways involved broad diverse cellular functions, namely in mitochondrial activity, protein kinase signaling, and immune function including autophagosome organization, exocytosis, receptor mediated endocytosis, phagocytic vesicle, secretory granule, and neutrophil degranulation.

### PANS cohort: Bulk whole blood RNA sequencing

We first performed whole blood bulk RNA sequencing in 20 children with PANS (mean age: 10.5 years (4-16), 62.5% males) versus 15 age and gender matched controls (mean age: 10.3 years (4-16), 60% males) (Supplementary Table 2 for patient details). Post RUV normalization, the principal component analysis of the bulk gene expression showed discrimination between the children with PANS versus controls (Figure 2B). There were 6914 differentially expressed genes (FDR <0.05), 3544 were upregulated (Figure 2C: in red) and 3369 were downregulated (Figure 2C: in blue).

Based on GSEA analysis using a ranked gene list, the top 5 upregulated Gene Ontology (GO) and Reactome pathways predominantly involved ribosomal biogenesis, translational processes, and RNA methyltransferase pathways (Figure 2D: in red, Supplementary Figure 7). In particular, the top three upregulated GO molecular function pathways were identified as structural constituents of ribosome, rRNA binding, and RNA methyltransferase activity (presented as CNET plot in Figure 3A: in red). Key genes that enriched the upregulated pathways included ribosomal genes associated with small subunit (40S) or large subunit (60S) of the eukaryotic and mitochondrial ribosome: *RPS, RPL, MRPS, MRPL*, as well as RNA and tRNA methyltransferase genes: *METTL, TRMT* (Figure 3B).

**Figure 3.**
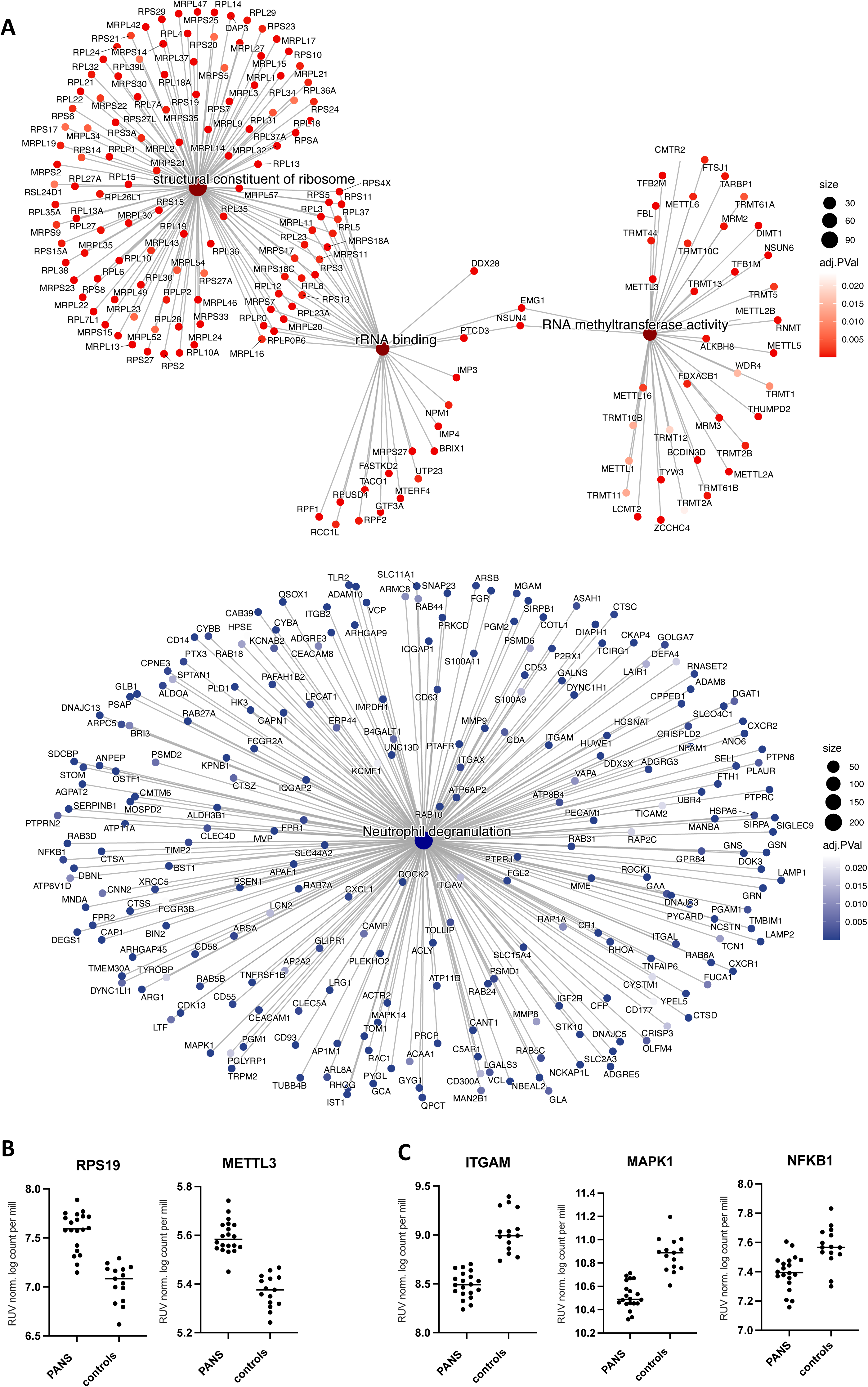
Connectivity network plots of top upregulated and downregulated bulk RNA sequencing in Paediatric acute neuropsychiatric syndrome patients, compared to controls. **(A)** Top panel: Connectivity network plot of the top 3 up-regulated enriched Gene Set Enrichment Analysis (GSEA) Gene Ontology molecular function (GO MF) pathways: structural constituent of ribosome, rRNA binding and RNA methyltransferase (in red). The enriched pathways are represented by their respective colors, and corresponding genes’ adjusted p value. Upregulated genes that enrich the pathways included ribosomal genes associated with small subunit (40S) or large subunit (60S) of the eukaryotic and mitochondrial ribosome: *RPS, RPL, MRPS, MRPL*, as well as RNA and tRNA methyltransferase genes: *METTL, TRMT*. Lower panel: Connectivity network (CNET) plot of the top down-regulated enriched GSEA Reactome pathway: neutrophil degranulation (in blue). The enriched pathway is represented by the respective colors, and corresponding genes’ adjusted p value. Downregulated genes that enrich the pathway included integrin genes: *ITGAM, ITGAL, ITGAX, ITGB2*, TLR genes: *TLR2, MAPK1, MAPK14, NFKB1*, cytokine/chemokine genes: *CXCR1, CXCR2, CXCL1, TNFRSF1B*, complement genes: *C5AR1*, and Fc-gamma receptor genes: *FCGR2A, FCGR3B*. **(B)** The y-axis depicts the log counts per million after the removal of unwanted variation (RUV) normalization, illustrating the upregulation of gene expression for representative genes *RPS19* and *METTL3* in PANS compared to controls. **(C)** The y-axis depicts the log counts per million after the removal of unwanted variation (RUV) normalization, illustrating the downregulation of gene expression for representative genes *ITGAM*, *MAPK1* and *NFKB1* in PANS compared to controls.

The top 5 downregulated GSEA GO and Reactome pathways involved diverse cellular functions, including mitochondrial activity, protein kinase signaling, and immune function involving autophagosome organization, exocytosis, receptor mediated endocytosis, phagocytic vesicle, secretory granule, and neutrophil degranulation (Figure 2D: in blue). The top downregulated Reactome pathway involved neutrophil degranulation (presented as CNET plot in Figure 3A: in blue). The corresponding downregulated genes included integrin genes: *ITGAM, ITGAL, ITGAX, ITGB2*, toll-like receptor (TLR) genes: *TLR2, MAPK1, MAPK14, NFKB1*, cytokine/chemokine genes: *CXCR1, CXCR2, CXCL1, TNFRSF1B*, complement genes: *C5AR1*, and Fc-gamma receptor genes: *FCGR2A, FCGR3B* (Figure 3C).

### PANS cohort: Single-cell blood RNA sequencing

Based on the findings from bulk RNA sequencing, we conducted a deeper investigation into cell-specific gene expression patterns through single-cell RNA sequencing. We performed single-cell RNA sequencing in two children with PANS (mean age: 8 years, 50% male, case summaries in Supplementary text), and compared them to 2 neurotypical controls (mean age: 11.5 years, 50% male).

Based on cell markers, we identified nine distinct cell types (Figure 4A). Adaptive immune cells, T cells (CD8 and CD4), and B cells constituted the largest proportion of cells across all samples (Figure 4B). Additionally, UMAP analysis of integrated biological samples revealed distinct cell clusters (Figure 4C). There were no significant differences observed in the distribution of cell clusters between individual PANS patients and controls (Supplementary Figure 8).

**Figure 4.**
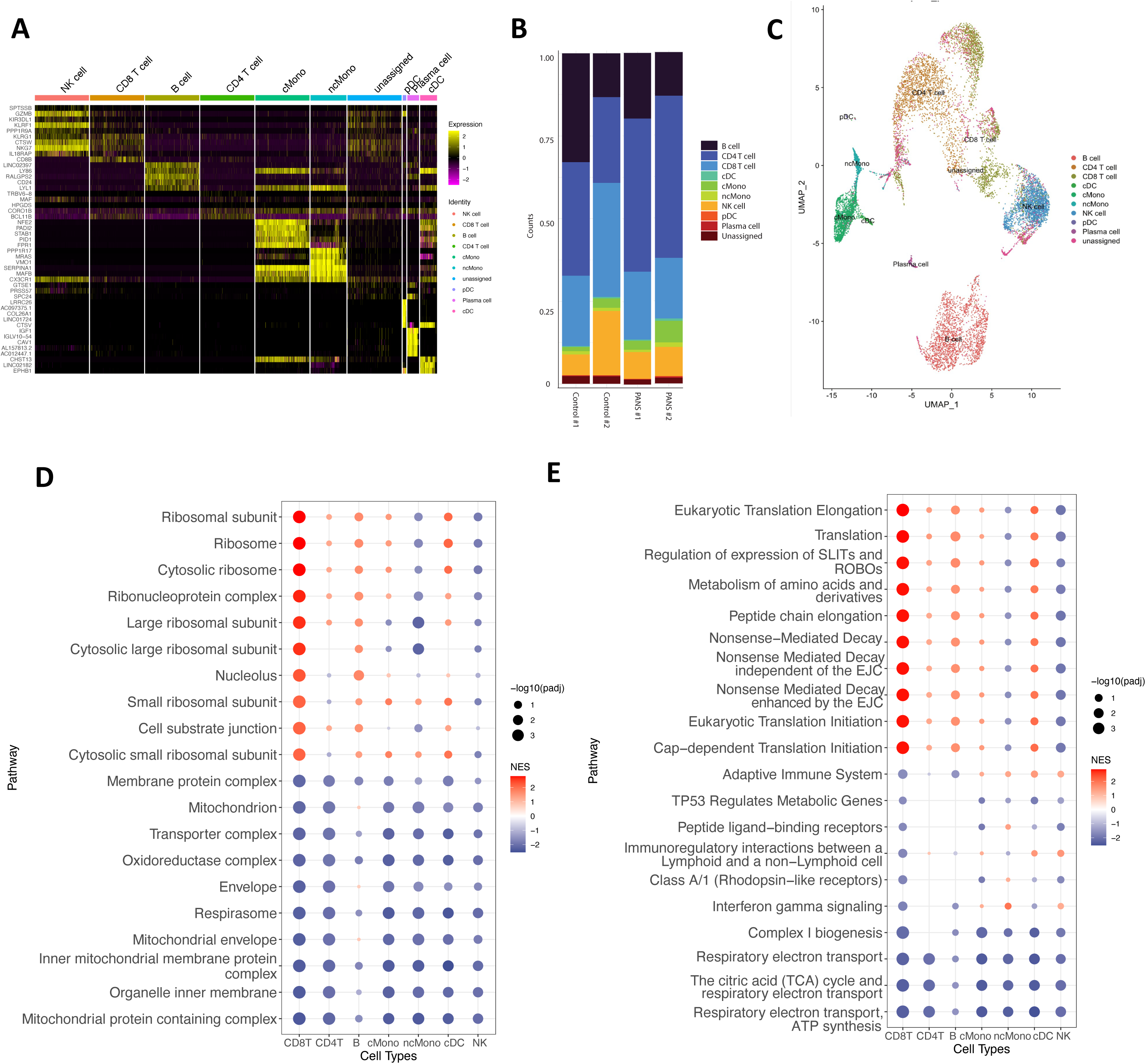
Single- cell blood RNA sequencing of Paediatric acute neuropsychiatric syndrome patients versus controls. **(A)** Heatmap of top differentially expressed genes based on cell type. Genes are the top markers of differential analysis between cell types (FDR <0.05) ranked by difference in proportion of cells expressing the respective gene. **(B)** Bar chart of cell population composition of the two controls and two PANS patient samples **(C)** Uniform manifold approximation and projection (UMAP) projection of all 4 samples, including 2 children with PANS and 2 control samples in single cell transcriptomics identified 9 unique clusters: NK cells, CD8 T cell, B cell, CD4 T cell, classical monocytes (cMono), non- classical monocytes (ncMono), plasmacytoid dendritic cell (pDC), classical dendritic cell (cDC). **(D)** Dot plot visualizing the top 30 GSEA GO cellular component pathways for CD8T cells. The dot’s colour represents the normalized enrichment score (NES), with red indicating upregulation, blue indicating downregulation, and zero represented by white. The size of each dot corresponds to the -log10(padj value) of the pathway. The NES scores of these pathways were mapped across all other cell types, demonstrating heterogeneity in cell type pathway enrichment. In terms of ribosome biogenesis pathways, it was found that adaptive immune cells exhibited upregulation, whereas the direction of these pathways was more varied in innate immune cells. In addition, cellular function, including mitochondrial activity, were downregulated across all cell types. **(E)** Dot plot visualizing the top 30 Reactome pathways for CD8T cells. The NES scores of these pathways were mapped across all other cell types, demonstrating heterogeneity in cell type pathway enrichment. In terms of translational pathways, it was found that adaptive immune cells exhibited upregulation, whereas the direction of these pathways was more varied in innate immune cells. Immune pathways were predominantly downregulated in adaptive immune cells and upregulated in innate immune cells.

The most significant enriched pathways were observed in CD8 T cells and B cells, likely attributed to their large population sizes in the sequencing data, indicating their substantial contribution to the identified pathway enrichments (Figure 4D, Figure 4E, Supplementary Figure 9, 10). GSEA cellular component (Figure 4D) and reactome analysis (Figure 4E) revealed predominantly upregulated pathways related to ribosome biogenesis and translation. Upon further investigation, ribosome biogenesis and translational pathways showed upregulation in adaptive immune cells, but the direction of these pathways was more varied in innate immune cells. Specifically, ribosomal and translational pathways were upregulated in classical monocytes and dendritic cells, but showed downregulation in non-classical monocytes and natural killer cells. Conversely, predominantly downregulated pathways included cellular pathways including mitochondrial, respiratory electron transport (Figure 4D and Figure 4E). Immune pathways including IFN-gamma signaling and adaptive immune system were predominantly downregulated in adaptive immune cells but upregulated in innate immune cells. Additionally, immune pathways including endocytic vesicle, phagocytic vesicle and secretory granule which were downregulated in bulk RNA sequencing, were upregulated in CD4 T cells and innate immune cells including classical monocytes, non-classical monocytes, and natural killer cells (Supplementary Figure 9, 10).

### PANS cohort: Toll like-receptor immune assay

Given the dysregulation of immune pathways uncovered by both bulk and single-cell RNA sequencing analyses, we aimed to determine the functional immune status of PBMCs in PANS. We assessed cytokine responsiveness upon stimulation of a critical innate immune pathway, the TLR pathway. TLR proteins expressed on immune cells recognize pathogen-associated molecular patterns like lipopolysaccharide (LPS), triggering a signaling cascade that produces pro-inflammatory cytokines and chemokines (Figure 5A). We measured the gene expression and cytokine production of LPS-stimulated PBMCs of 7 children with PANS (mean age 10.6 years (range 5-15), 57.1% male) versus 7 neurotypical controls (mean age 11.9 years (range 8-15), 57.1% male) (Fig. 5B).

**Figure 5.**
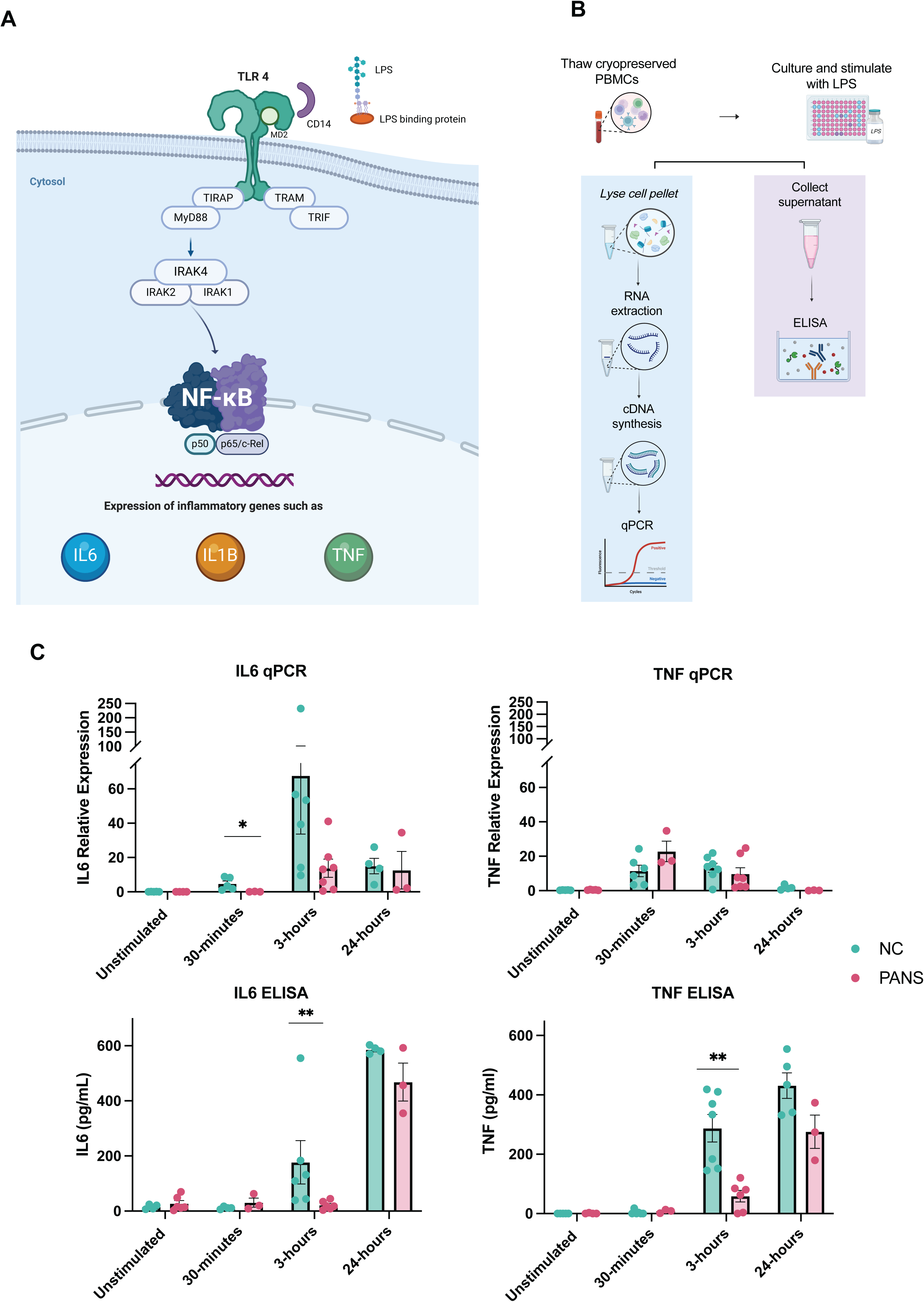
Toll-like receptor stimulation analysis in Paediatric acute neuropsychiatric syndrome patients compared to controls. **(A)** The Toll-like receptor (TLR) pathway is a crucial component of the innate immune system responsible for detecting and responding to various pathogens. TLR proteins expressed on immune cells recognize pathogen-associated molecular patterns like lipopolysaccharide (LPS), triggering a signaling cascade that produces pro-inflammatory cytokines, such as interleukin (IL)6 and tumour necrosis factor (TNF). TLR4= Toll-like receptor 4, TIRAP= Toll/interleukin-1 receptor (TIR) domain-containing adaptor protein, TRIF= TIR-domain-containing adaptor-inducing interferon-**beta**, TRAM= TRIF-related adaptor molecule, MyD88= myeloid differentiation primary response protein 88, IRAK= interleukin-1 receptor associated kinase, NF-κB (nuclear factor kappa light chain enhancer of activated B cells) **(B)** Gene expression and cytokine production of LPS stimulated peripheral blood mononuclear cells (PBMCs) of PANS patients compared to controls. PBMCs of PANS and controls were stimulated with 500ng/mL LPS for 30 minutes, 3 hours, and 24 hours. Gene expression of IL6, and TNF were measured using quantitative reverse transcription polymerase chain reaction. Target gene expression was normalized to household gene, Beta-2-Microglobulin. In addition, release of IL6 and TNF into the cell culture media was measured using ELISA. **(C)** In PBMCs stimulated with LPS, IL6 gene (RNA) expression was lower at both 30 minutes (p < 0.05) and 3 hours (p < 0.05) in PANS compared to controls, and IL6 and TNF protein production (ELISA) was significantly reduced at 3 hours (p < 0.01) in PANS compared to controls (Figure 5C). Bars and whiskers represent median with interquartile range; n = 7. Mann-Whitney test, *P < 0·05, **P < 0·01.

In PBMCs stimulated with LPS, *IL6* expression was reduced at both 30 minutes (p < 0.05) and 3 hours (p < 0.05) in PANS compared to controls, and IL6 and TNF (protein) was reduced at 3 hours (p < 0.01) in PANS compared to controls (Figure 5C). This indicated a reduced responsiveness to LPS stimulation in PBMCs of PANS compared to controls.

### PANS IVIg cohort: Clinical response to IVIg treatment

Within the PANS cohort, we observed a clinical pattern of response to IVIg in 9 children who received open-label 4 weekly IVIg. Although the therapeutic response to IVIg varied, the effect commenced early (often a few days after infusion), but only lasted for around two to three weeks, and the beneficial effect often waned before the next infusion (Figure 6A). Most of our patients required chronic monthly use of IVIg due to the ongoing nature of their symptoms. For example, in one patient, we observed gradual improvement of OCD symptoms with monthly IVIg treatment over 6 months, however OCD symptoms appeared to worsen after stopping IVIg, requiring re-commencement (Figure 6B).

**Figure 6.**
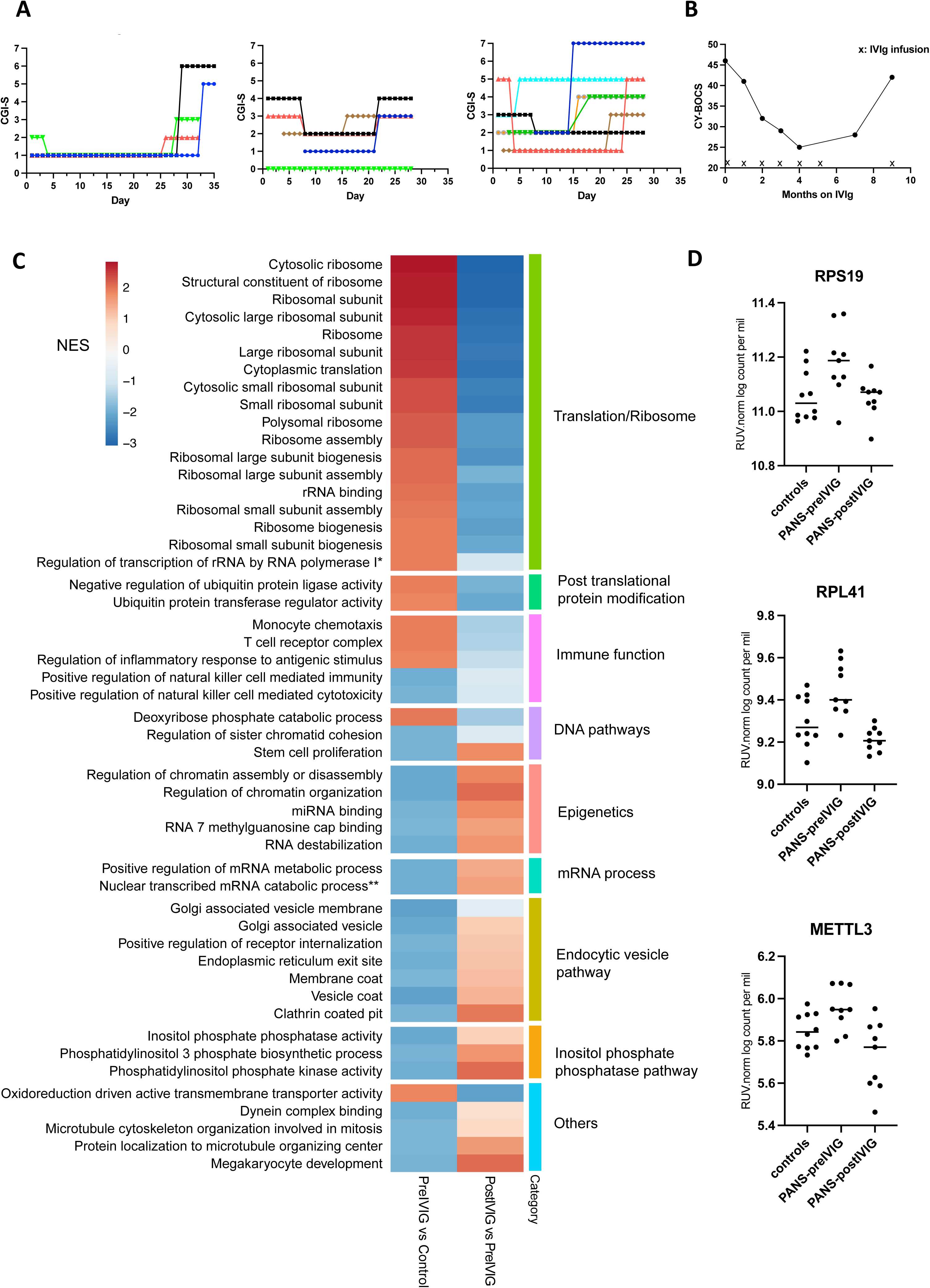
Bulk blood RNA sequencing of children with Paediatric acute-onset neuropsychiatric syndrome (PANS) before and after intravenous immunoglobulin (IVIg) treatment, compared to controls. **(A)** Clinical response of three children with PANS who received IVIg was prospectively assessed using the Clinical Global Impression Scale- Severity (CGI-S) to document the therapeutic response to IVIg . We charted their daily CGI-S scores over 4-8 four-week cycles (different colour represents a separate 4-week cycle). CGI-S followed a seven-point scale where 1 is normal and 7 is extremely ill. The clinical benefit of IVIg is typically seen 1-7 days after infusion and lasts for 2-3 weeks, and then the clinical benefit wanes. **(B)** Clinical response (OCD symptoms) of a child with PANS who received monthly IVIg treatment over 6 months using the Children’s Yale-Brown Obsessive Compulsive Scale (CY- BOCS) score. CY-BOCS score has a total severity score of 0-50 to chart the progress of OCD symptoms. IVIg infusions are marked with X. We observed gradual improvement in OCD symptoms this child over 6 months. However, OCD symptoms were observed to relapse after stopping IVIg, requiring re-commencement of IVIg. **(C)** We compared the normalized enrichment scores (NES) for the top 50 Gene Set Enrichment Analysis (GSEA) Gene Ontology (GO) pathways present in: (1) PANS pre IVIg versus control (left column), and (2) PANS post IVIg versus PANS pre IVIg (right column). Pathways upregulated (red) in PANS pre-IVIg versus controls including ribosomal biogenesis, translational, post-translational protein modifications, and immune pathways, were downregulated (blue) post-IVIg treatment. Pathways that were downregulated in the PANS pre-IVIG versus controls including immune pathways, epigenetics, and broad cellular function, showed upregulation after IVIg treatment. *Regulation of transcription of nucleolar large rRNA by RNA polymerase I **Nuclear transcribed mRNA catabolic process deadenylation-dependent decay **(D)** The expression of representative genes *RPS19, RPL41*, and *METTL3* was upregulated in PANS Pre IVIg compared to controls. After IVIg treatment, the gene expression of these genes was decreased, comparable to levels in controls. The log counts per million post-removal of unwanted variation (RUV) normalization are plotted on the y-axis.

### PANS IVIg cohort: Bulk whole blood RNA sequencing

Given the abnormalities found in gene translation and cellular function at baseline in PANS, we next explored the mechanistic action of IVIg in PANS. We performed bulk RNA sequencing in 9 children with PANS (mean age: 10.1 years (7-15), 55.6% males) before and after receiving IVIg treatment compared to 10 healthy controls (mean age: 10.8 years (8-17), 50% males). Following RUV normalization, principal component analysis (PCA) of the bulk RNA sequencing data revealed robust discrimination between children with PANS at baseline and controls. Additionally, PANS patients post-IVIg treatment appeared to exhibit a closer proximity to the control group, suggesting a potential therapeutic effect of IVIg treatment on RNA sequencing profiles in PANS patients (Supplementary Figure 11). There were 541 differentially expressed genes (FDR <0.05) in PANS at baseline versus controls, and 4200 differentially expressed genes (FDR <0.05) in PANS post IVIg treatment compared to pre IVIg treatment.

We compared the normalized enrichment scores (NES) for the top 50 GSEA GO pathways present in: (1) PANS pre IVIg versus control (left column), and (2) PANS post IVIg versus PANS pre IVIg (right column) (Figure 6C). We were able to reproduce our baseline RNA sequencing findings in PANS versus controls in this second (replication) PANS cohort. In PANS pre IVIG versus control, the most upregulated pathways involved ribosomal function and translation, whereas the most downregulated genes involved immune function, endocytosis and cell signalling (Figure 6C). The pathways that were upregulated in PANS pre IVIg treatment were downregulated post IVIg treatment (ribosomal biogenesis, translational, post translational protein modifications). Pathways that were downregulated in the PANS pre IVIg treatment were upregulated post IVIg treatment (immune, epigenetic, and cellular function) (Figure 6C, Figure 6D, Supplementary Figure 12).

## Discussion

The prevailing paradigm regarding the pathogenesis of NDDs recognizes a complex interplay between genetic, epigenetic, and environmental factors. Environmental influences at critical windows of vulnerability have been shown to affect fetal brain development in animal models^27–29^. Proposed mechanisms of early-life programming in neurodevelopment involve epigenetic ‘priming’ of microglial and peripheral immune cells, resulting in systemic immune dysregulation^30–32^. A ‘two-hit’ model explains how the prenatal disruptions in brain development increases offspring susceptibility to recurrent infections and abnormal neurodevelopmental trajectories^27, 29^. While prenatal first ‘hits’, such as maternal inflammation during pregnancy, have been firmly linked to NDD risk, the connection between postnatal ‘hits’ and NDD symptoms in humans remains less defined^27^.

Brain development is a highly dynamic process, involving a fine orchestration of neurological functions. Within this context, episodic “regressions” in developmental skills have been previously described and diagnosed as autistic regression or PANS^11, 13, 14^. In our cohort, children with NDDs unsurprisingly exhibited a high degree of emotional and behavioural issues. Notably, we also observed a high frequency of loss of developmental skills, often triggered by infection or stress, in children with NDD compared to controls. This observation suggests that loss of developmental skills may be a common occurrence in NDDs, and not exclusive to autistic regression or PANS. We observed increased reported childhood infections and allergic conditions in children with NDDs compared to controls, consistent with previous studies^9, 33–35^. The underlying mechanisms behind the association between recurrent infections and neurodevelopmental or psychiatric manifestations is poorly understood. To better delineate disease pathways underlying this association, we focussed our investigation on children fulfilling PANS criteria. Our hypothesis was that PANS is not an autoimmune encephalitis, but instead a gene regulatory disorder associated with peripheral and brain immune dysregulation.

We showed that CSF examination of children with PANS is typically normal. We were unable to identify evidence of neuroinflammation as seen in autoimmune encephalitis in our PANS patients, although subtle changes in CSF are sometimes seen in PANS, such as mildly elevated CSF protein^36^. In addition, routine immune testing in PANS revealed normal quantitative immune cells in peripheral blood, and no evidence of systemic inflammation. In a clinical cohort, while brain cell RNA sequencing is not possible, peripheral blood immune analysis is feasible. In this study, children experienced recurrent infections and exhibited abnormal immune responses, making peripheral blood immune cell transcriptomics highly relevant in understanding their condition. Key findings from our blood RNA-sequencing analyses of PANS patients included upregulation of translational pathways enriched with ribosomal proteins, and downregulated pathways of broad cellular function and immune pathways. The translation of information encoded by mRNA into functional proteins occurs in ribosomes, which plays critical roles in brain development^37^. There are established associations between disruption of translational machinery and NDDs. Firstly, monogenic mutations affecting components of the translational process, such as ribosomal proteins or translational factors can cause NDDs^38–40^. Secondly, environmental exposures like infections and stressors can disrupt translational processes by impacting ribosomal biogenesis^41^. Ribosomal biogenesis is critical for maintaining homeostasis during stress such as proteostasis imbalance, nutrient deprivation, and oxidative stress^41^. Perturbations to ribosomal biogenesis can lead to reduction of protein synthesis crucial for dendritic and synaptic function, contributing to changes in neuronal connectivity associated with NDDs^40^. In our study, we observed a large number of upregulated *RPL* and *RPS* genes coding for 40S and 60S ribosomal subunits in PANS versus controls, also seen in in blood leukocytes and post-mortem cortical tissue of individuals with ASD^42^. Consistent findings across these studies supports the notion that ribosomal protein dysregulation in both brain and peripheral blood is a central feature in the pathophysiology of NDDs in general, and not specific to PANS ^42^.

We hypothesize that early life environmental factors alter the epigenetic regulation of immune cells in NDDs, resulting in disturbance to the translational processes via ribosomal biogenesis. We found epigenetic pathways including RNA methyltransferase and chromatin organization to be dysregulated in children with PANS. Recent studies highlight the environmental impact on epigenetics and epitranscriptomics, which play pivotal roles in regulating translational processes^43, 44^. Histone methyltransferases (*KMT)*, histone demethylase (*KDM)*, and RNA methyltransferases (*METTL*), play crucial roles in regulating ribosomal gene transcription^45–48^. Modifications to histone lysine methylation have been shown to modulate ribosomal expression via post translational modifications, aiding cell adaptation to stressful environmental conditions^49^. In addition, ribosomal proteins also have non-ribosomal functions, such as immune functions^50^. Ribosomal stress results in the release and accumulation of ribosome-free ribosomal proteins that are involved in immune signaling pathways, which may also be contributing to downstream immune dysregulation in PANS^50^. However, other interactions between immune, translational, and epigenetic processes are also plausible. Immune cells are highly adaptable to environmental stimuli, such as infections, and their responses involve complex interactions with epigenetic mechanisms and ribosomal biogenesis. For example, T cell activation, marked by the release of effector molecules, can initiate reprogramming of cellular metabolism, typically accompanied by increased ribosomal biogenesis and enhanced activity of RNA cap methyltransferases^51, 52^. Thus, it is possible that our RNA sequencing data reflects activation of certain immune cell populations, in early life or in response to recent environmental stimuli. Nevertheless, the precise interactions between epigenetic, translational, and immune mechanisms warrants further investigations in the context of PANS.

This study observed a diverse downregulation of cellular functions using RNA sequencing in PANS, encompassing mitochondrial activity, receptor-mediated endocytosis, exocytosis, and immune pathways. These findings are consistent with RNA sequencing studies of blood and brain samples of other NDDs^53, 54^. Specifically, immune factors are increasingly recognised to operate at the gene-environment interface during NDD pathogenesis. Integrated brain transcriptome and epigenetic analyses of individuals with NDD demonstrate convergent dysregulated immune pathways^55, 56^. Abnormal peripheral immune responses, including abnormal cytokine production, immunoglobulin levels, and altered cellular response to stimuli, have also been reported in individuals with NDDs ^57^. We identified a downregulated immune signal in blood RNA sequencing, implicating various innate immune pathways, enriched in genes related to TLR, integrin, complement, cytokine, and chemokine signaling. The cytokine hypo-responsiveness to stimulation via LPS in the children with PANS in our study provides additional evidence of innate immune dysfunction in PANS. While this study primarily highlights innate immune dysregulation, our single-cell RNA sequencing also reveals immune dysregulation in adaptive immune cells, such as CD8+ T cells and B cells. Although TLR is primarily associated with innate immune responses, TLR also has direct and indirect effects of T cells function^58^. Given the fact that we used a PBMC fraction, further studies should explore differential TLR response in individual cell types, such as monocytes and lymphocytes. Further immunological testing on both innate and adaptive immune cells is needed to determine whether this dysregulation is global or restricted to specific cell types. We propose that children with PANS have global suppression of immune function, with downstream deficits in cell debris clearance, and potential secondary inflammation. This finding correlates with the recurrent infections in children with NDDs that we and others have observed^10, 59^.

Early-life exposures to environmental factors, can lead to ‘epigenetic’ programming, impacting immune cells both peripherally and in the brain^60, 61^. There is a dynamic cross-talk between microglia and other myeloid-derived cells^62^. While our study has revealed peripheral blood immune dysregulation in PANS, it is plausible that microglia also have impaired function in individuals with PANS. Microglia are crucial for immune surveillance, regulating neuronal functions, and influencing synaptic connections through synaptic pruning^6, 63^. Disruption to microglia-mediated synaptic pruning is a key process in NDDs and psychiatric disorders ^63^. Previous positron emission tomography imaging studies of individuals with PANS revealed reactive microglial states^64^. Animal models of maternal immune activation show perturbations in offspring microglia function, including impaired phagocytosis, resulting in aberrant proinflammatory states, contributing to neurobehavioral abnormalities^65^. We propose that early life environmentally-driven epigenetic changes to the neuroimmune axis can result in neuroimmune dysfunction, contributing to neurodevelopmental and psychiatric disorders ^62, 66^.

The use of immunotherapy in PANS, particularly IVIg is reported to be clinically beneficial, but its effectiveness is controversial and its immunomodulatory effects remain unknown ^16, 17, 67^. IVIg has multiple mechanisms of action in inflammatory diseases, including modulation of expression of Fc receptors, interference with complement activation and cytokines, and regulation of immune cell activation^68^. Our study is the first to provide insights on the biological effects of IVIg in PANS. Firstly, we found that IVIg modified dysregulated pathways involving both innate and adaptive immunity present at baseline in PANS, demonstrating the broad immunomodulatory effects of IVIg in PANS. Secondly, IVIg appeared to modify other key dysregulated pathways in PANS related to ribosomal biogenesis, translational, and post-translational protein modifications, as well as epigenetic pathways. Although studies on the effects of IVIg on the translational program are limited, other mechanisms affecting gene regulation, via methylation and microRNA alterations, have been reported in Kawasaki disease and immunodeficiency^69, 70^. Thirdly, we found upregulated pathways in inositol phosphate phosphatase activity post IVIg treatment in PANS. The effects of IVIg on the Fc gamma receptor via inositol phosphate phosphatase pathways have also been demonstrated in immune thrombocytopenia purpura ^71^. It should be emphasized that although we describe the benefit of IVIg in some individuals with PANS and demonstrate potential biological effects of IVIg on gene regulation, this does not provide proof of effect. Robust randomised-controlled clinical trials with significant clinical benefits are needed to provide evidence for IVIg in PANS. Further investigations into the impacts of IVIg on peripheral-brain immune crosstalk, along with identifying gene signatures to predict individual treatment responsiveness to IVIg are crucial for personalizing treatment options for individuals with PANS. Additionally, correlating clinical responsiveness to IVIg with biological changes in gene expression, will help to delineate key pathways critical for alleviating PANS symptoms. Further comparisons with biological investigations in children receiving IVIg for other indications, such as hypogammaglobulinemia or autoimmune conditions (i.e. immune thrombocytopenic purpura etc) could clarify whether the observed changes in the patients with PANS are unique to PANS or represent a broader response to IVIg. In vitro cell-based studies are also needed to identify specific mechanisms of IVIg in PANS, distinguishing them from general effects of IVIg on blood cells. Studying biological effects in patients with hypogammaglobulinemia, who typically receive a lower dose of IVIg, may also help to establish a potential dose-response relationship. Lastly, exploring oral treatments with sustained effects is important considering the short-lived clinical response to IVIg.

In this study, we demonstrate a common gene regulatory signature in PANS, replicated across two cohorts using bulk RNA sequencing and further confirmed in two patients through single- cell RNA sequencing. We propose that this signature arises from the interaction between environmental factors and genetic vulnerability, mediated by the epigenetic machinery. Nonetheless, we acknowledge the possible influence of DNA variants on this signature. We did not undertake comprehensive genomic DNA testing in our cohort, as this was not the focus of our study, and acknowledge that detection of rare, highly penetrant, DNA variation is uncommon in PANS and NDD outside of intellectual disability. Of the 9 children with PANS and mild intellectual disability in our cohort, comparative genomic hybridization (CGH) was performed in 9 and whole exome sequencing/gene panel in 7, and was negative in all. Limitations of our study include the retrospective nature of the childhood infection and loss of skills history. The high rates of childhood infections and loss of skills may be partly related to patient enrolment from a specialized NDD clinic, which may be influenced by retrospective recall bias, and referral bias. Additionally, controlling for complex human variables and potential confounders was challenging. We were unable to control for socioeconomic status, which is an important confounder as lower socioeconomic status seen in our NDD cohort could be associated with higher rates of childhood infections and other allergic diseases. Second, our omics analysis involved a small sample size with considerable heterogeneity in symptomology and phase of disease. Although some of the children with PANS were in the chronic phase of disease, all of them had ongoing debilitating neuropsychiatric symptoms, requiring psychotherapy and psychiatric medications. Longitudinal RNA sequencing at different time points in PANS (acute and chronic phase) will help us better understand the stability of the gene expression signature. Additionally, while there may be persistent gene expression changes in people with chronic infections, we ensured that the children with PANS did not have significant acute or uncontrolled chronic infections 4 weeks before blood taking. Gaining appropriate controls was challenging, half of our controls were being investigated for growth or puberty delay. Thirdly, patients were on conventional psychiatric medications which could potentially affect gene expression; however subgroup analysis of PANS patients did not reveal significant differences in gene expression between patients receiving psychiatric medications versus those who were not (data not shown). Fourth, it is important to note that bulk RNA sequencing of whole blood includes all leukocyte populations, whereas 10X Genomics single-cell RNA sequencing is based on PBMCs (excluding neutrophils). Fifth, our current study is based on peripheral blood and not brain, as brain is not a feasible tissue to investigate in humans. However, we hypothesize that peripheral blood cells are representative of brain cells, given the fact that putative environmental drivers of immune signature have effects across organs with different cellular origins. We propose that RNA sequencing signatures in peripheral blood will transpire to be a valuable diagnostic tool in NDDs^72^. Further studies to examine blood and brain RNA signatures in animal models are needed to further explore the blood-brain correlation.

Larger prospective cohort studies are needed, combining environmental exposures, omics analyses (DNA, RNA and epigenetic modifications) with detailed immune function testing, to unpack gene-environment interactions in PANS compared to the broader NDD spectrum. In our PANS cohort, a significant proportion of children had pre-existing NDDs. Although PANS commonly occurs in previously healthy children, emerging clinical evidence indicates that the constellation of PANS symptoms can also manifest in children with pre-existing NDDs, with an overlapping genetic background observed between PANS and NDDs^3^. In this study, we have not done direct comparisons of biological investigations in PANS compared to other non-PANS NDDs (ASD, ADHD, OCD, Tourette syndrome), and this comparison is important in future studies. Future studies including subgroups of patients with PANS and NDDs will be critical to determine if PANS is a subgroup with a unique biological mechanism, or if children with PANS represent the more severe end of NDD spectrum with similar genetic, environmental and epigenetic underpinnings. These efforts aim to advance precision medicine in diagnostics, prognostication, and therapeutics for NDDs.

In conclusion, we propose that the pathogenesis of PANS involves epigenetic dysregulation which results in downstream immune dysregulation ^53^. We propose that PANS is not a discrete ‘autoimmune’ entity but represents an important clinical phenotype of NDDs centred around epigenetics. Epigenetic and immune modulating therapies require further investigation in PANS and other NDDs ^73, 74^.

## Data availability

All relevant data are included in the manuscript or supplemental material. Bulk RNA sequencing and single-cell RNA sequencing data has been uploaded to Gene Expression Omnibus (GEO) under accession GSE278680. Raw data files and R code are available to be shared upon request to the corresponding author

## Acknowledgements

We are grateful for the study participants who contributed to this research. We would like to thank the doctors and nurses in the Department of Paediatric Endocrinology at The Children’s Hospital at Westmead, Sydney for their help and support to recruit controls. We would like to thank Children’s Medical Research Institute and Australian Genome Research Facility for providing expertise in sequencing and bioinformatic support. We would like to thank PANS Australia, Stepping Stones Therapy for Children, and Dr Leila Masson for their kind donations for this study.

## Author contributions

VH, SA, SP and RD designed the research studies. VH, BK, SY, HN, HJ, IP, SM, RD recruited the patients and performed clinical assessments. VH, SA, BK, MG, EM, JY, SB, KK, ET, PL, RD conducted experiments and acquired and analyzed the data. MH, MG, FB, WG, SP and RD oversaw the experiments and data analysis. VH, SA, BG, NA, ML, EM performed computational analyses. VH and SA drafted the manuscript and shared responsibilities as co-first authors. The order of the co-first author’s names was determined by workload. SP and RD critically revised the manuscript and shared responsibilities as senior authors.

## Supplementary material

Supplementary Text

Supplementary Table

Supplementary Figure

