## SupplementaryMaterials for "Epigenetic, ribosomal, and immune dysregulation in Paediatric Acute-Onset Neuropsychiatric Syndrome"

### Slide 1
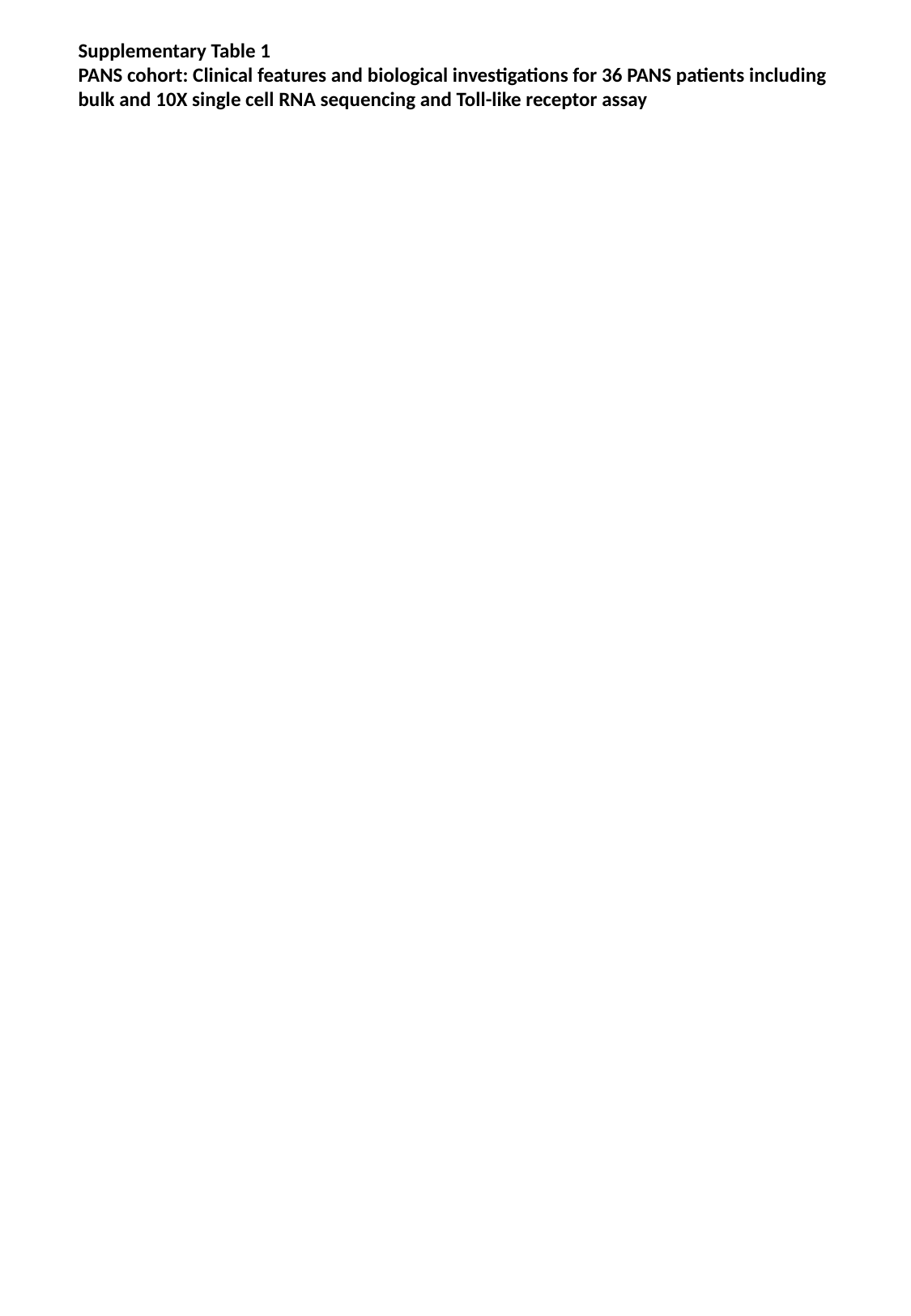

Supplementary Table 1
PANS cohort: Clinical features and biological investigations for 36 PANS patients including
bulk and 10X single cell RNA sequencing and Toll-like receptor assay

### Slide 2
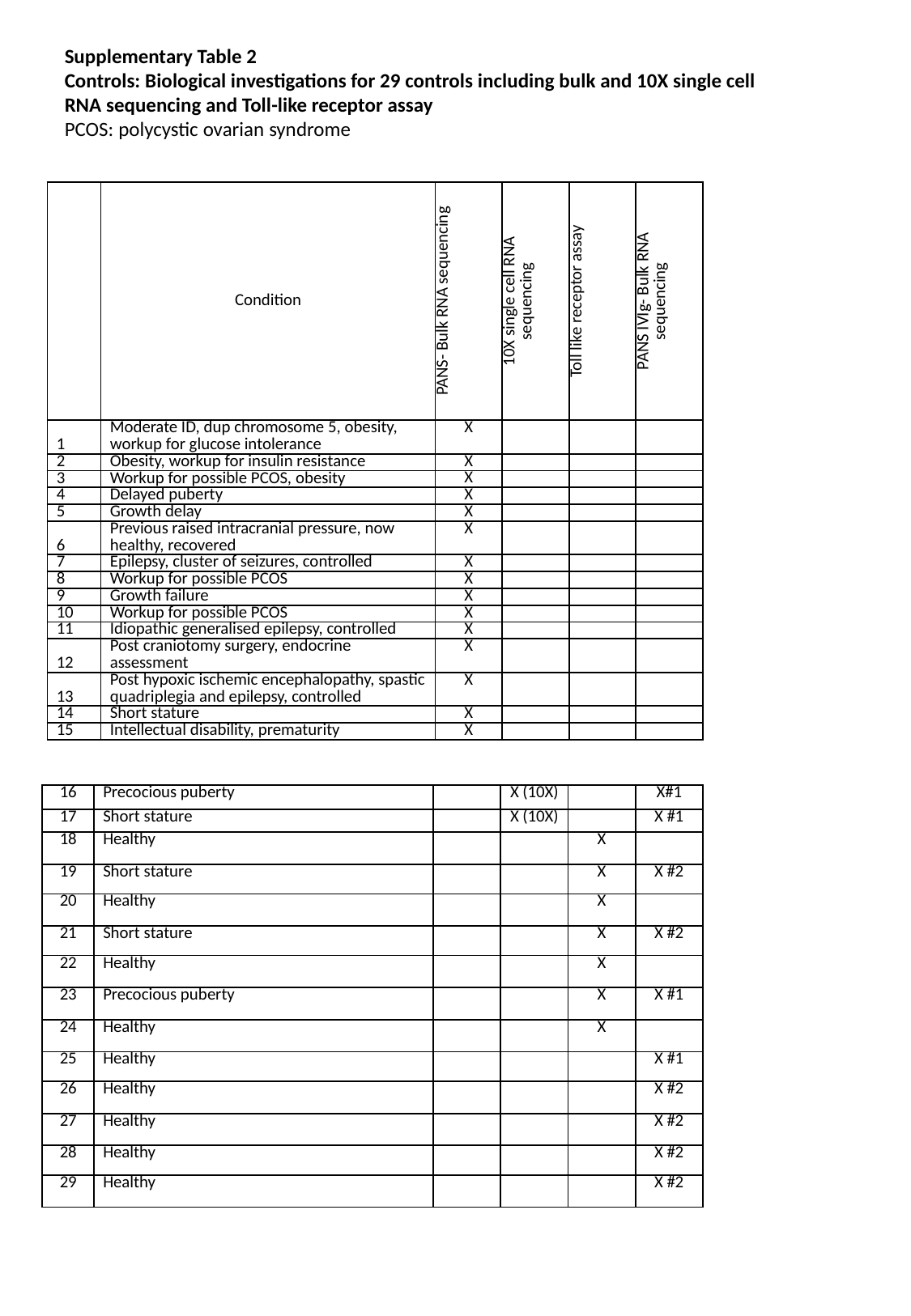

Supplementary Table 2
Controls: Biological investigations for 29 controls including bulk and 10X single cell
RNA sequencing and Toll-like receptor assay
PCOS: polycystic ovarian syndrome
| | Condition | PANS- Bulk RNA sequencing | 10X single cell RNA sequencing | Toll like receptor assay | PANS IVIg- Bulk RNA sequencing |
| --- | --- | --- | --- | --- | --- |
| 1 | Moderate ID, dup chromosome 5, obesity, workup for glucose intolerance | X | | | |
| 2 | Obesity, workup for insulin resistance | X | | | |
| 3 | Workup for possible PCOS, obesity | X | | | |
| 4 | Delayed puberty | X | | | |
| 5 | Growth delay | X | | | |
| 6 | Previous raised intracranial pressure, now healthy, recovered | X | | | |
| 7 | Epilepsy, cluster of seizures, controlled | X | | | |
| 8 | Workup for possible PCOS | X | | | |
| 9 | Growth failure | X | | | |
| 10 | Workup for possible PCOS | X | | | |
| 11 | Idiopathic generalised epilepsy, controlled | X | | | |
| 12 | Post craniotomy surgery, endocrine assessment | X | | | |
| 13 | Post hypoxic ischemic encephalopathy, spastic quadriplegia and epilepsy, controlled | X | | | |
| 14 | Short stature | X | | | |
| 15 | Intellectual disability, prematurity | X | | | |
| 16 | Precocious puberty | | X (10X) | | X#1 |
| --- | --- | --- | --- | --- | --- |
| 17 | Short stature | | X (10X) | | X #1 |
| 18 | Healthy | | | X | |
| 19 | Short stature | | | X | X #2 |
| 20 | Healthy | | | X | |
| 21 | Short stature | | | X | X #2 |
| 22 | Healthy | | | X | |
| 23 | Precocious puberty | | | X | X #1 |
| 24 | Healthy | | | X | |
| 25 | Healthy | | | | X #1 |
| 26 | Healthy | | | | X #2 |
| 27 | Healthy | | | | X #2 |
| 28 | Healthy | | | | X #2 |
| 29 | Healthy | | | | X #2 |

### Slide 3
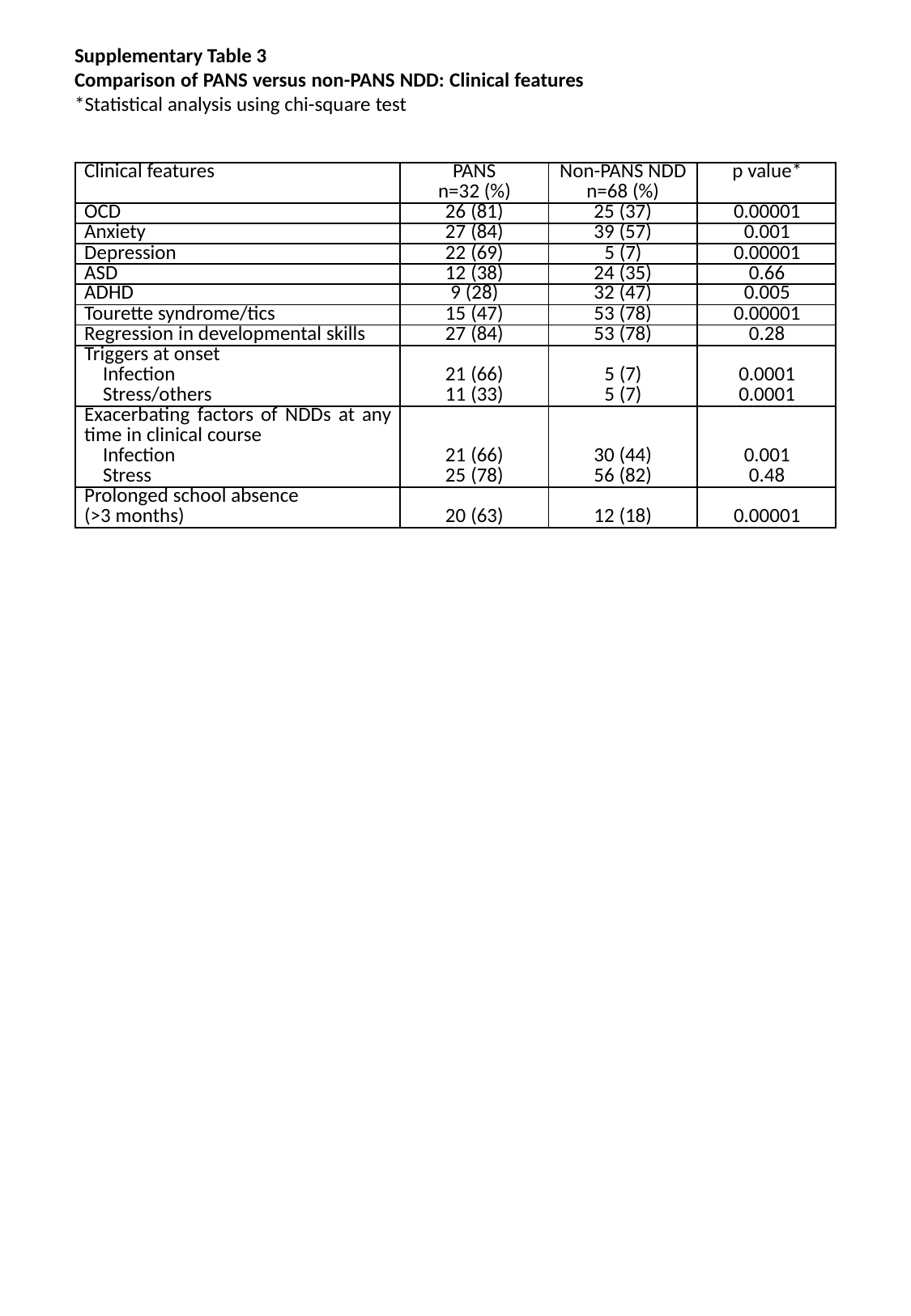

Supplementary Table 3
Comparison of PANS versus non-PANS NDD: Clinical features
*Statistical analysis using chi-square test
| Clinical features | PANS n=32 (%) | Non-PANS NDD n=68 (%) | p value\* |
| --- | --- | --- | --- |
| OCD | 26 (81) | 25 (37) | 0.00001 |
| Anxiety | 27 (84) | 39 (57) | 0.001 |
| Depression | 22 (69) | 5 (7) | 0.00001 |
| ASD | 12 (38) | 24 (35) | 0.66 |
| ADHD | 9 (28) | 32 (47) | 0.005 |
| Tourette syndrome/tics | 15 (47) | 53 (78) | 0.00001 |
| Regression in developmental skills | 27 (84) | 53 (78) | 0.28 |
| Triggers at onset Infection Stress/others | 21 (66) 11 (33) | 5 (7) 5 (7) | 0.0001 0.0001 |
| Exacerbating factors of NDDs at any time in clinical course Infection Stress | 21 (66) 25 (78) | 30 (44) 56 (82) | 0.001 0.48 |
| Prolonged school absence (>3 months) | 20 (63) | 12 (18) | 0.00001 |

### Slide 4
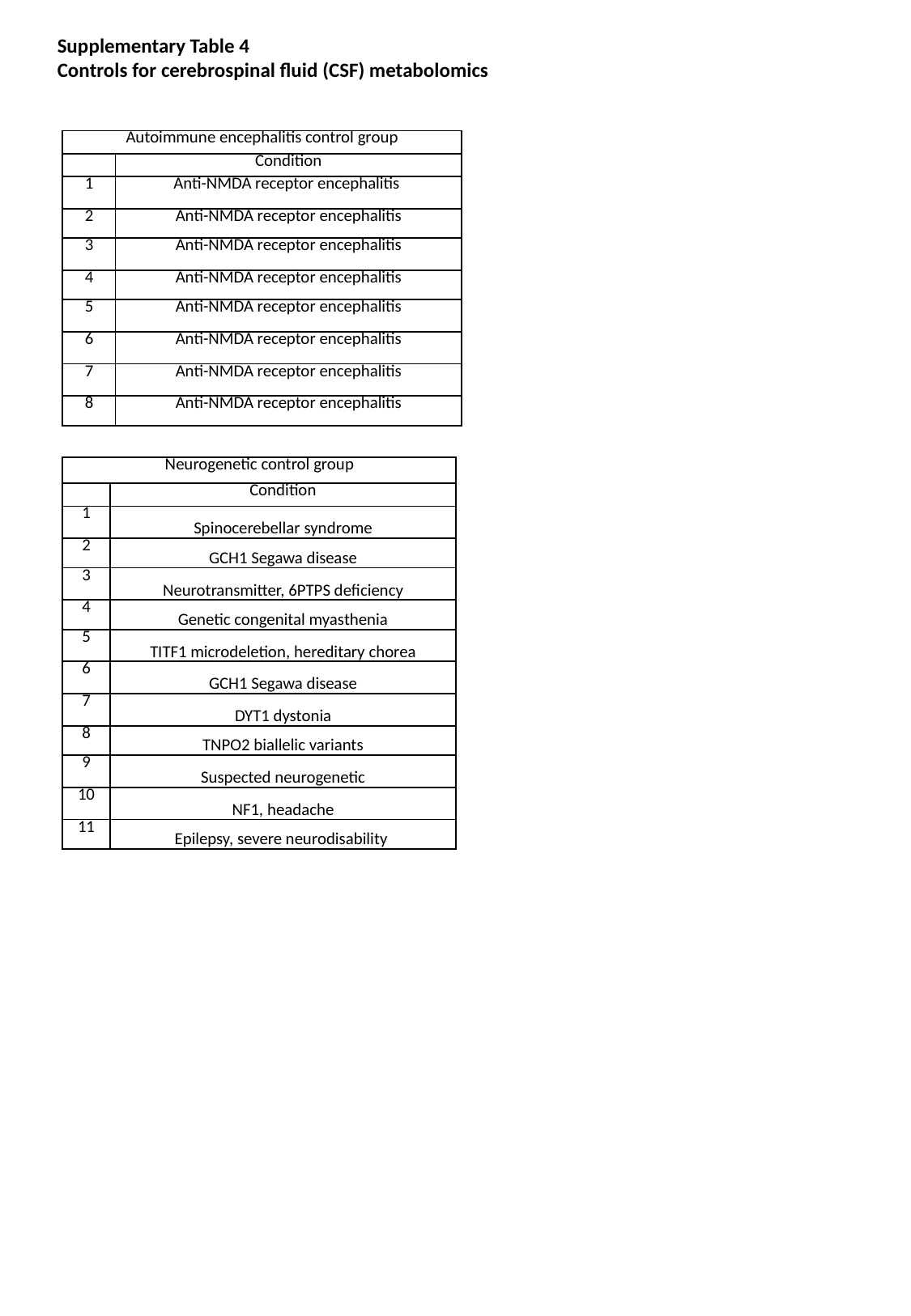

Supplementary Table 4
Controls for cerebrospinal fluid (CSF) metabolomics
| Autoimmune encephalitis control group | |
| --- | --- |
| | Condition |
| 1 | Anti-NMDA receptor encephalitis |
| 2 | Anti-NMDA receptor encephalitis |
| 3 | Anti-NMDA receptor encephalitis |
| 4 | Anti-NMDA receptor encephalitis |
| 5 | Anti-NMDA receptor encephalitis |
| 6 | Anti-NMDA receptor encephalitis |
| 7 | Anti-NMDA receptor encephalitis |
| 8 | Anti-NMDA receptor encephalitis |
| Neurogenetic control group | |
| --- | --- |
| | Condition |
| 1 | Spinocerebellar syndrome |
| 2 | GCH1 Segawa disease |
| 3 | Neurotransmitter, 6PTPS deficiency |
| 4 | Genetic congenital myasthenia |
| 5 | TITF1 microdeletion, hereditary chorea |
| 6 | GCH1 Segawa disease |
| 7 | DYT1 dystonia |
| 8 | TNPO2 biallelic variants |
| 9 | Suspected neurogenetic |
| 10 | NF1, headache |
| 11 | Epilepsy, severe neurodisability |

### Slide 5
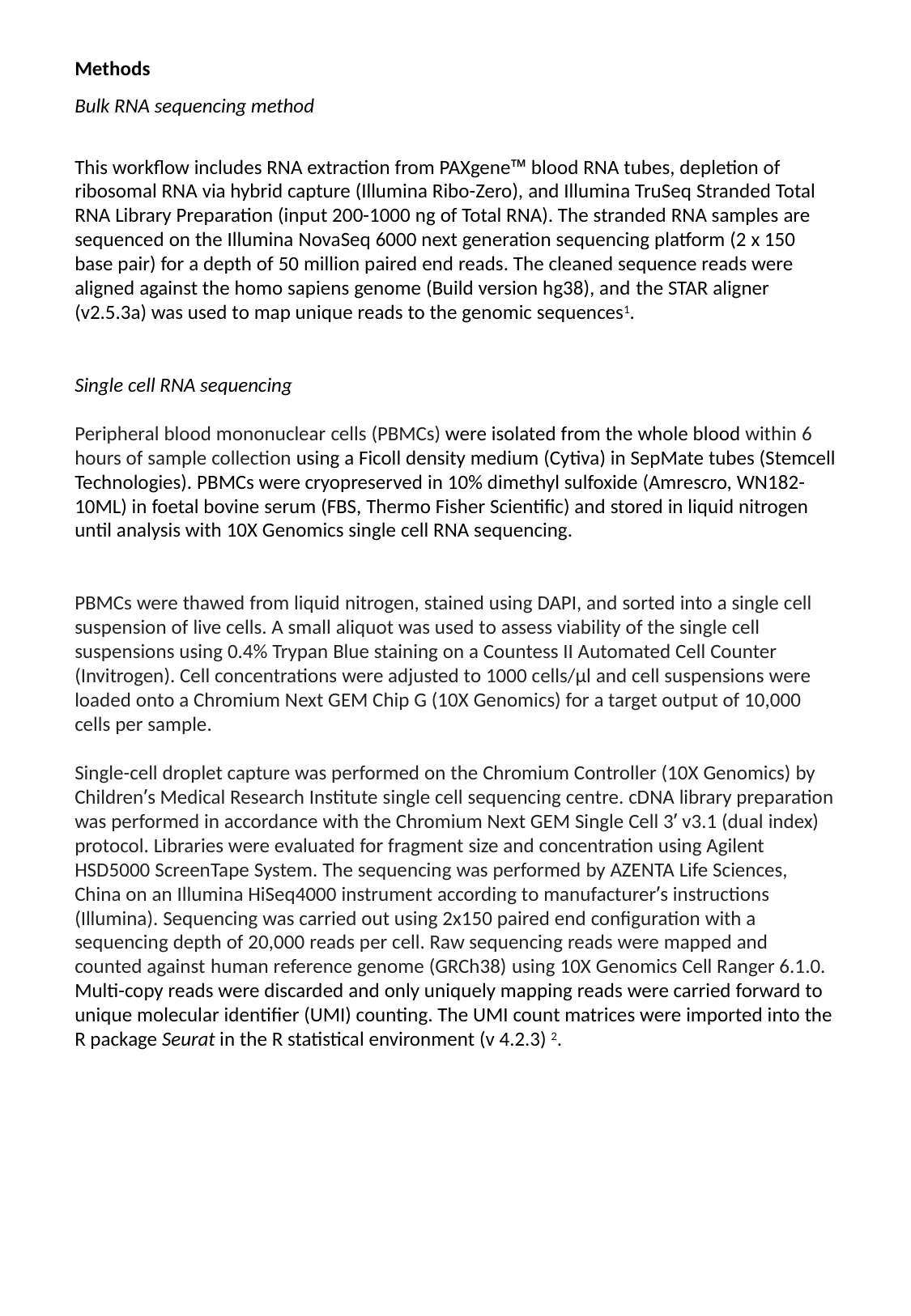

Methods
Bulk RNA sequencing method
This workflow includes RNA extraction from PAXgene™ blood RNA tubes, depletion of ribosomal RNA via hybrid capture (Illumina Ribo-Zero), and Illumina TruSeq Stranded Total RNA Library Preparation (input 200-1000 ng of Total RNA). The stranded RNA samples are sequenced on the Illumina NovaSeq 6000 next generation sequencing platform (2 x 150 base pair) for a depth of 50 million paired end reads. The cleaned sequence reads were aligned against the homo sapiens genome (Build version hg38), and the STAR aligner (v2.5.3a) was used to map unique reads to the genomic sequences^1^.
Single cell RNA sequencing
Peripheral blood mononuclear cells (PBMCs) were isolated from the whole blood within 6 hours of sample collection using a Ficoll density medium (Cytiva) in SepMate tubes (Stemcell Technologies). PBMCs were cryopreserved in 10% dimethyl sulfoxide (Amrescro, WN182-10ML) in foetal bovine serum (FBS, Thermo Fisher Scientific) and stored in liquid nitrogen until analysis with 10X Genomics single cell RNA sequencing.
PBMCs were thawed from liquid nitrogen, stained using DAPI, and sorted into a single cell suspension of live cells. A small aliquot was used to assess viability of the single cell suspensions using 0.4% Trypan Blue staining on a Countess II Automated Cell Counter (Invitrogen). Cell concentrations were adjusted to 1000 cells/μl and cell suspensions were loaded onto a Chromium Next GEM Chip G (10X Genomics) for a target output of 10,000 cells per sample.
Single-cell droplet capture was performed on the Chromium Controller (10X Genomics) by Children’s Medical Research Institute single cell sequencing centre. cDNA library preparation was performed in accordance with the Chromium Next GEM Single Cell 3’ v3.1 (dual index) protocol. Libraries were evaluated for fragment size and concentration using Agilent HSD5000 ScreenTape System. The sequencing was performed by AZENTA Life Sciences, China on an Illumina HiSeq4000 instrument according to manufacturer’s instructions (Illumina). Sequencing was carried out using 2x150 paired end configuration with a sequencing depth of 20,000 reads per cell. Raw sequencing reads were mapped and counted against human reference genome (GRCh38) using 10X Genomics Cell Ranger 6.1.0. Multi-copy reads were discarded and only uniquely mapping reads were carried forward to unique molecular identifier (UMI) counting. The UMI count matrices were imported into the R package Seurat in the R statistical environment (v 4.2.3) ^2^.

### Slide 6
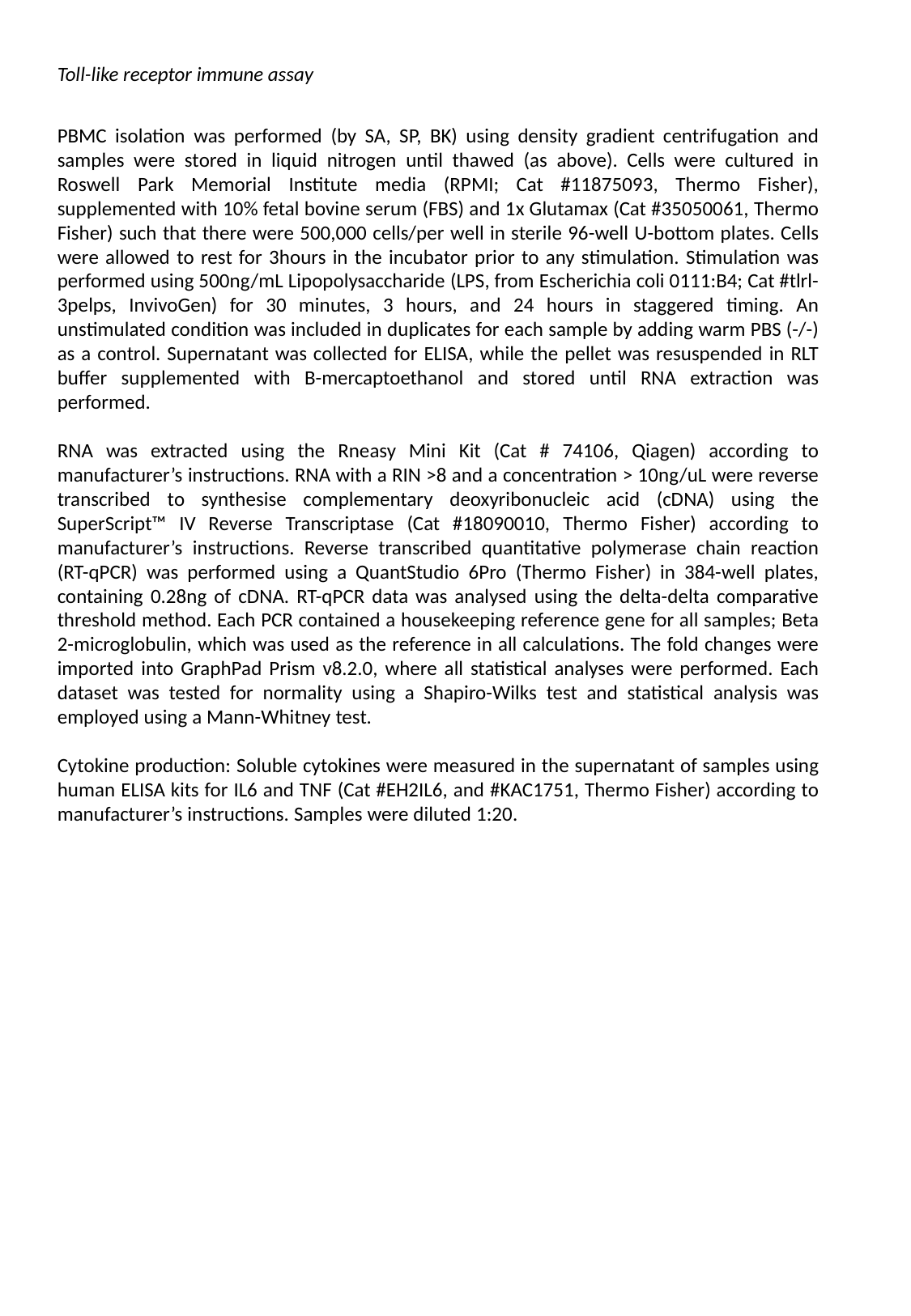

Toll-like receptor immune assay
PBMC isolation was performed (by SA, SP, BK) using density gradient centrifugation and samples were stored in liquid nitrogen until thawed (as above). Cells were cultured in Roswell Park Memorial Institute media (RPMI; Cat #11875093, Thermo Fisher), supplemented with 10% fetal bovine serum (FBS) and 1x Glutamax (Cat #35050061, Thermo Fisher) such that there were 500,000 cells/per well in sterile 96-well U-bottom plates. Cells were allowed to rest for 3hours in the incubator prior to any stimulation. Stimulation was performed using 500ng/mL Lipopolysaccharide (LPS, from Escherichia coli 0111:B4; Cat #tlrl-3pelps, InvivoGen) for 30 minutes, 3 hours, and 24 hours in staggered timing. An unstimulated condition was included in duplicates for each sample by adding warm PBS (-/-) as a control. Supernatant was collected for ELISA, while the pellet was resuspended in RLT buffer supplemented with B-mercaptoethanol and stored until RNA extraction was performed.
RNA was extracted using the Rneasy Mini Kit (Cat # 74106, Qiagen) according to manufacturer’s instructions. RNA with a RIN >8 and a concentration > 10ng/uL were reverse transcribed to synthesise complementary deoxyribonucleic acid (cDNA) using the SuperScript™ IV Reverse Transcriptase (Cat #18090010, Thermo Fisher) according to manufacturer’s instructions. Reverse transcribed quantitative polymerase chain reaction (RT-qPCR) was performed using a QuantStudio 6Pro (Thermo Fisher) in 384-well plates, containing 0.28ng of cDNA. RT-qPCR data was analysed using the delta-delta comparative threshold method. Each PCR contained a housekeeping reference gene for all samples; Beta 2-microglobulin, which was used as the reference in all calculations. The fold changes were imported into GraphPad Prism v8.2.0, where all statistical analyses were performed. Each dataset was tested for normality using a Shapiro-Wilks test and statistical analysis was employed using a Mann-Whitney test.
Cytokine production: Soluble cytokines were measured in the supernatant of samples using human ELISA kits for IL6 and TNF (Cat #EH2IL6, and #KAC1751, Thermo Fisher) according to manufacturer’s instructions. Samples were diluted 1:20.

### Slide 7
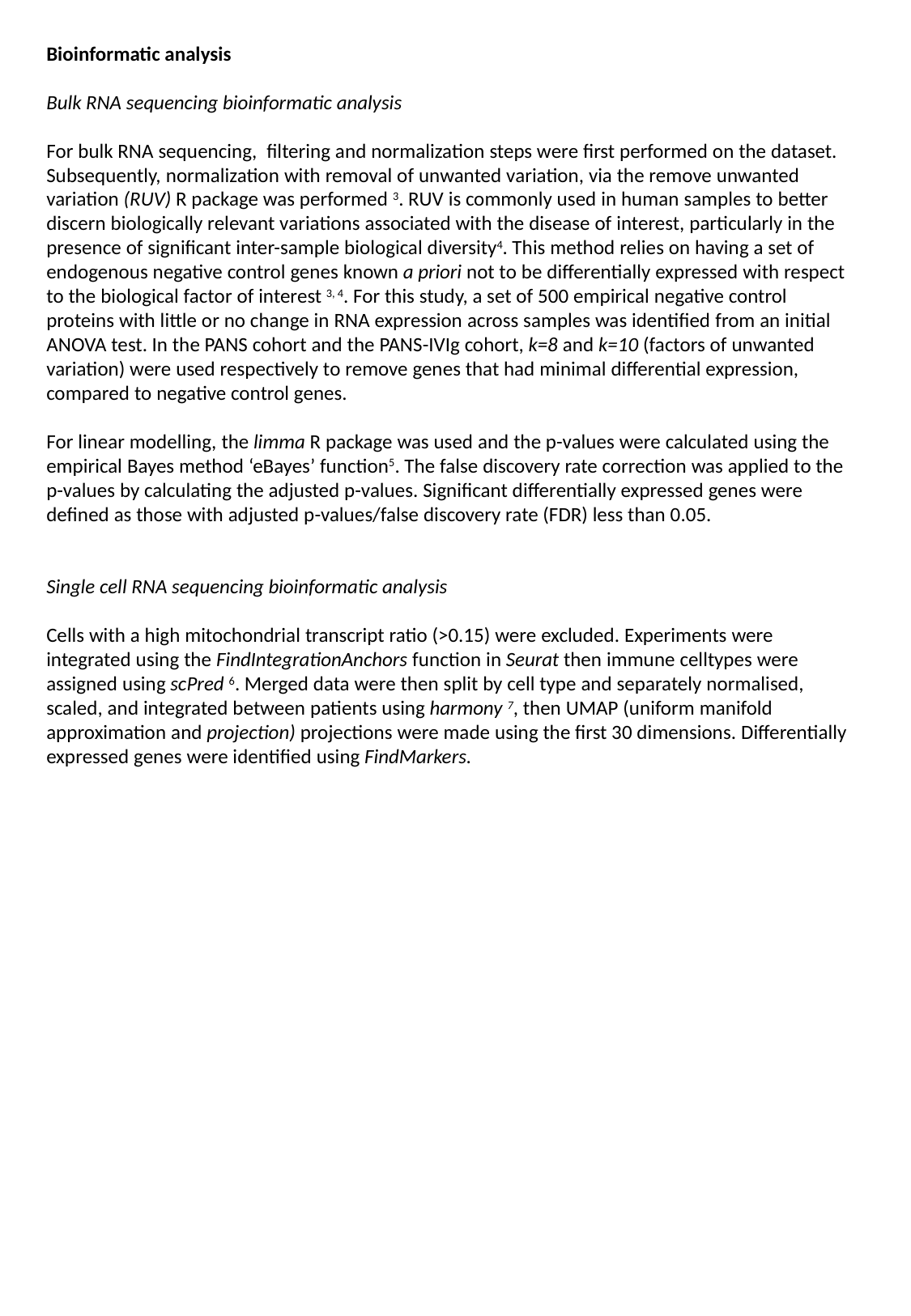

Bioinformatic analysis
Bulk RNA sequencing bioinformatic analysis
For bulk RNA sequencing, filtering and normalization steps were first performed on the dataset. Subsequently, normalization with removal of unwanted variation, via the remove unwanted variation (RUV) R package was performed ^3^. RUV is commonly used in human samples to better discern biologically relevant variations associated with the disease of interest, particularly in the presence of significant inter-sample biological diversity^4^. This method relies on having a set of endogenous negative control genes known a priori not to be differentially expressed with respect to the biological factor of interest ^3, 4^. For this study, a set of 500 empirical negative control proteins with little or no change in RNA expression across samples was identified from an initial ANOVA test. In the PANS cohort and the PANS-IVIg cohort, k=8 and k=10 (factors of unwanted variation) were used respectively to remove genes that had minimal differential expression, compared to negative control genes.
For linear modelling, the limma R package was used and the p-values were calculated using the empirical Bayes method ‘eBayes’ function^5^. The false discovery rate correction was applied to the p-values by calculating the adjusted p-values. Significant differentially expressed genes were defined as those with adjusted p-values/false discovery rate (FDR) less than 0.05.
Single cell RNA sequencing bioinformatic analysis
Cells with a high mitochondrial transcript ratio (>0.15) were excluded. Experiments were integrated using the FindIntegrationAnchors function in Seurat then immune celltypes were assigned using scPred ^6^. Merged data were then split by cell type and separately normalised, scaled, and integrated between patients using harmony ^7^, then UMAP (uniform manifold approximation and projection) projections were made using the first 30 dimensions. Differentially expressed genes were identified using FindMarkers.

### Slide 8
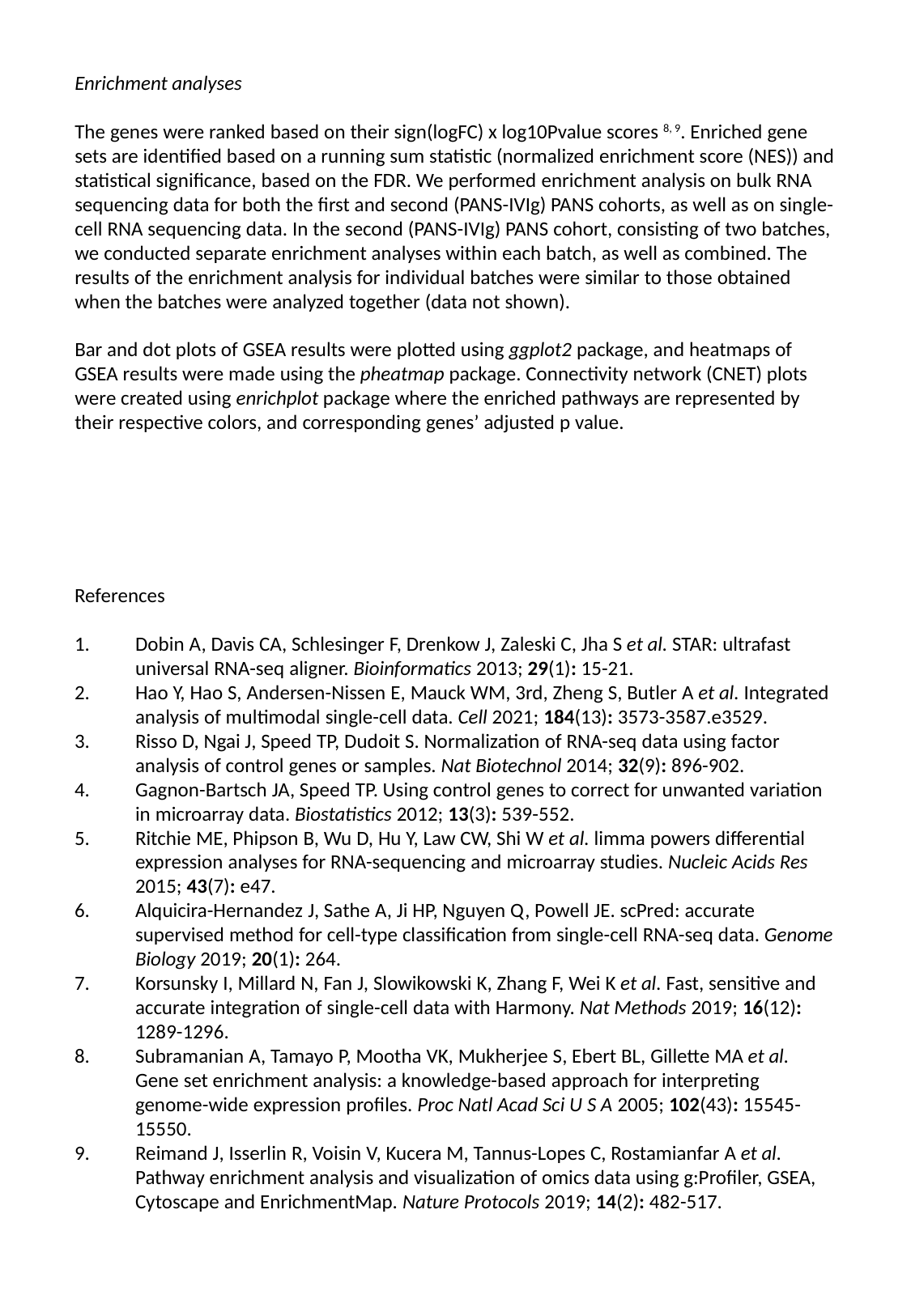

Enrichment analyses
The genes were ranked based on their sign(logFC) x log10Pvalue scores ^8, 9^. Enriched gene sets are identified based on a running sum statistic (normalized enrichment score (NES)) and statistical significance, based on the FDR. We performed enrichment analysis on bulk RNA sequencing data for both the first and second (PANS-IVIg) PANS cohorts, as well as on single-cell RNA sequencing data. In the second (PANS-IVIg) PANS cohort, consisting of two batches, we conducted separate enrichment analyses within each batch, as well as combined. The results of the enrichment analysis for individual batches were similar to those obtained when the batches were analyzed together (data not shown).
Bar and dot plots of GSEA results were plotted using ggplot2 package, and heatmaps of GSEA results were made using the pheatmap package. Connectivity network (CNET) plots were created using enrichplot package where the enriched pathways are represented by their respective colors, and corresponding genes’ adjusted p value.
References
1.	Dobin A, Davis CA, Schlesinger F, Drenkow J, Zaleski C, Jha S et al. STAR: ultrafast universal RNA-seq aligner. Bioinformatics 2013; 29(1): 15-21.
2.	Hao Y, Hao S, Andersen-Nissen E, Mauck WM, 3rd, Zheng S, Butler A et al. Integrated analysis of multimodal single-cell data. Cell 2021; 184(13): 3573-3587.e3529.
3.	Risso D, Ngai J, Speed TP, Dudoit S. Normalization of RNA-seq data using factor analysis of control genes or samples. Nat Biotechnol 2014; 32(9): 896-902.
4.	Gagnon-Bartsch JA, Speed TP. Using control genes to correct for unwanted variation in microarray data. Biostatistics 2012; 13(3): 539-552.
5.	Ritchie ME, Phipson B, Wu D, Hu Y, Law CW, Shi W et al. limma powers differential expression analyses for RNA-sequencing and microarray studies. Nucleic Acids Res 2015; 43(7): e47.
6.	Alquicira-Hernandez J, Sathe A, Ji HP, Nguyen Q, Powell JE. scPred: accurate supervised method for cell-type classification from single-cell RNA-seq data. Genome Biology 2019; 20(1): 264.
7.	Korsunsky I, Millard N, Fan J, Slowikowski K, Zhang F, Wei K et al. Fast, sensitive and accurate integration of single-cell data with Harmony. Nat Methods 2019; 16(12): 1289-1296.
8.	Subramanian A, Tamayo P, Mootha VK, Mukherjee S, Ebert BL, Gillette MA et al. Gene set enrichment analysis: a knowledge-based approach for interpreting genome-wide expression profiles. Proc Natl Acad Sci U S A 2005; 102(43): 15545-15550.
9.	Reimand J, Isserlin R, Voisin V, Kucera M, Tannus-Lopes C, Rostamianfar A et al. Pathway enrichment analysis and visualization of omics data using g:Profiler, GSEA, Cytoscape and EnrichmentMap. Nature Protocols 2019; 14(2): 482-517.

### Slide 9
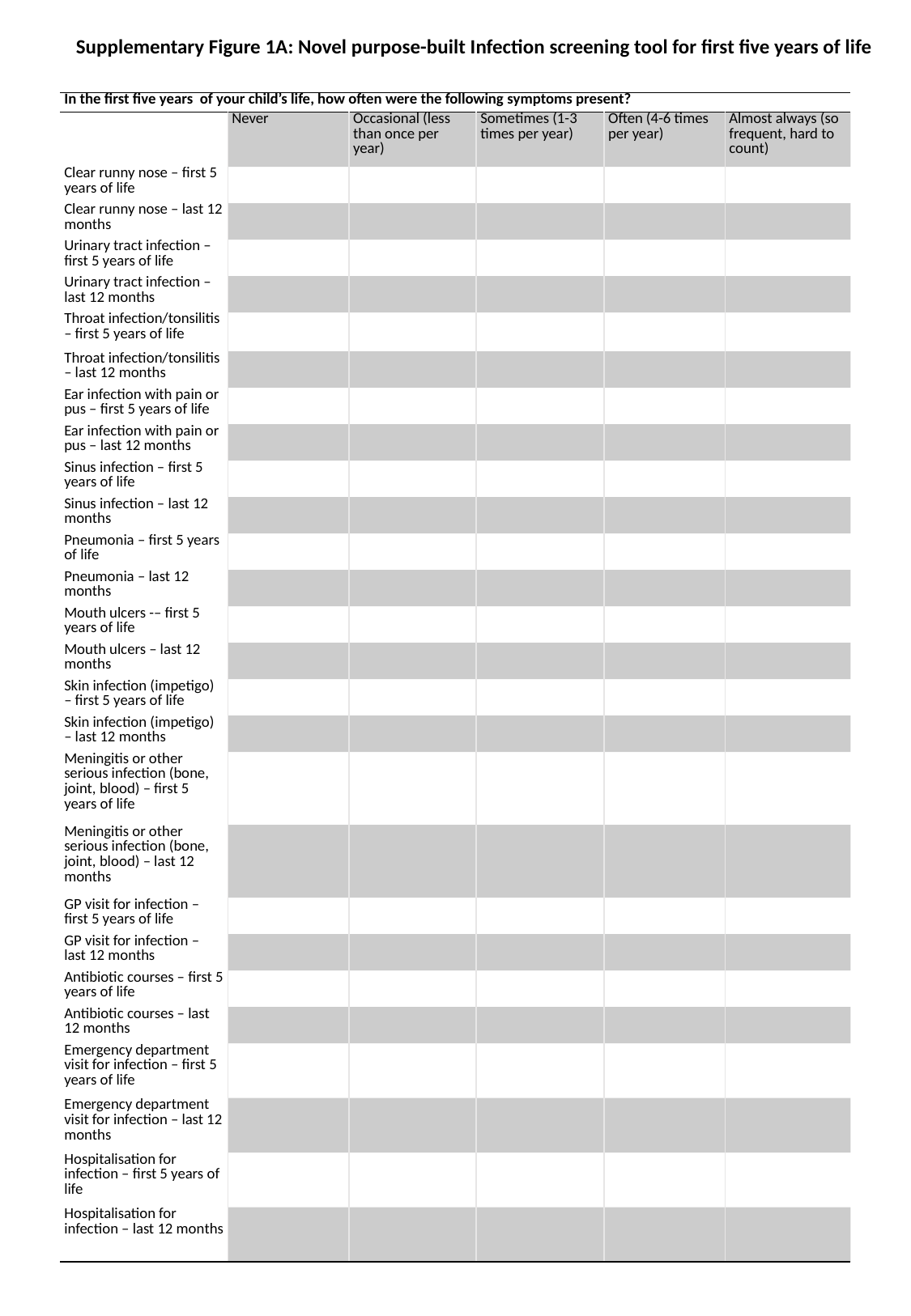

Supplementary Figure 1A: Novel purpose-built Infection screening tool for first five years of life
| In the first five years of your child’s life, how often were the following symptoms present? | | | | | |
| --- | --- | --- | --- | --- | --- |
| | Never | Occasional (less than once per year) | Sometimes (1-3 times per year) | Often (4-6 times per year) | Almost always (so frequent, hard to count) |
| Clear runny nose – first 5 years of life | | | | | |
| Clear runny nose – last 12 months | | | | | |
| Urinary tract infection – first 5 years of life | | | | | |
| Urinary tract infection – last 12 months | | | | | |
| Throat infection/tonsilitis – first 5 years of life | | | | | |
| Throat infection/tonsilitis – last 12 months | | | | | |
| Ear infection with pain or pus – first 5 years of life | | | | | |
| Ear infection with pain or pus – last 12 months | | | | | |
| Sinus infection – first 5 years of life | | | | | |
| Sinus infection – last 12 months | | | | | |
| Pneumonia – first 5 years of life | | | | | |
| Pneumonia – last 12 months | | | | | |
| Mouth ulcers -– first 5 years of life | | | | | |
| Mouth ulcers – last 12 months | | | | | |
| Skin infection (impetigo) – first 5 years of life | | | | | |
| Skin infection (impetigo) – last 12 months | | | | | |
| Meningitis or other serious infection (bone, joint, blood) – first 5 years of life | | | | | |
| Meningitis or other serious infection (bone, joint, blood) – last 12 months | | | | | |
| GP visit for infection – first 5 years of life | | | | | |
| GP visit for infection – last 12 months | | | | | |
| Antibiotic courses – first 5 years of life | | | | | |
| Antibiotic courses – last 12 months | | | | | |
| Emergency department visit for infection – first 5 years of life | | | | | |
| Emergency department visit for infection – last 12 months | | | | | |
| Hospitalisation for infection – first 5 years of life | | | | | |
| Hospitalisation for infection – last 12 months | | | | | |

### Slide 10
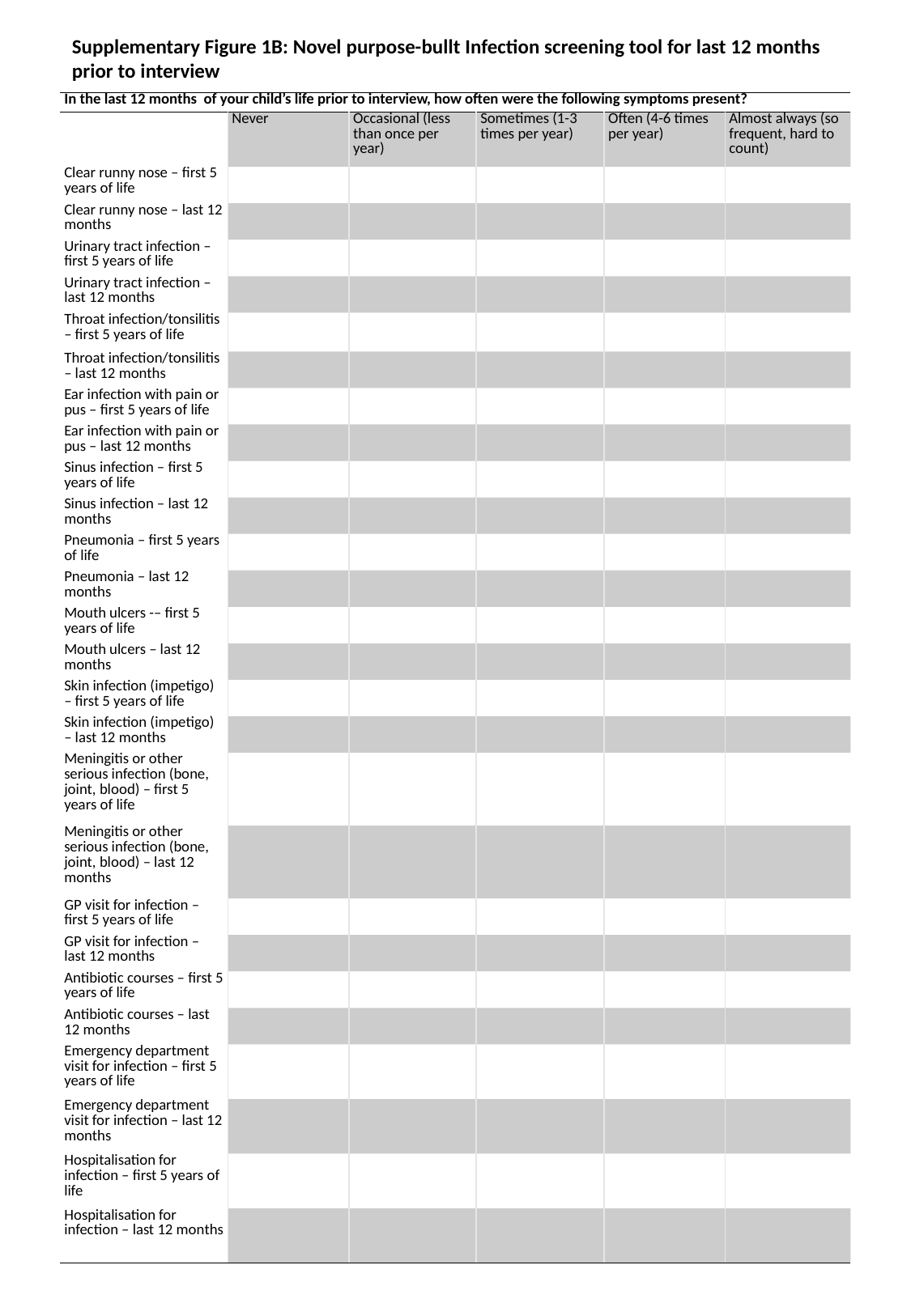

Supplementary Figure 1B: Novel purpose-bullt Infection screening tool for last 12 months prior to interview
| In the last 12 months of your child’s life prior to interview, how often were the following symptoms present? | | | | | |
| --- | --- | --- | --- | --- | --- |
| | Never | Occasional (less than once per year) | Sometimes (1-3 times per year) | Often (4-6 times per year) | Almost always (so frequent, hard to count) |
| Clear runny nose – first 5 years of life | | | | | |
| Clear runny nose – last 12 months | | | | | |
| Urinary tract infection – first 5 years of life | | | | | |
| Urinary tract infection – last 12 months | | | | | |
| Throat infection/tonsilitis – first 5 years of life | | | | | |
| Throat infection/tonsilitis – last 12 months | | | | | |
| Ear infection with pain or pus – first 5 years of life | | | | | |
| Ear infection with pain or pus – last 12 months | | | | | |
| Sinus infection – first 5 years of life | | | | | |
| Sinus infection – last 12 months | | | | | |
| Pneumonia – first 5 years of life | | | | | |
| Pneumonia – last 12 months | | | | | |
| Mouth ulcers -– first 5 years of life | | | | | |
| Mouth ulcers – last 12 months | | | | | |
| Skin infection (impetigo) – first 5 years of life | | | | | |
| Skin infection (impetigo) – last 12 months | | | | | |
| Meningitis or other serious infection (bone, joint, blood) – first 5 years of life | | | | | |
| Meningitis or other serious infection (bone, joint, blood) – last 12 months | | | | | |
| GP visit for infection – first 5 years of life | | | | | |
| GP visit for infection – last 12 months | | | | | |
| Antibiotic courses – first 5 years of life | | | | | |
| Antibiotic courses – last 12 months | | | | | |
| Emergency department visit for infection – first 5 years of life | | | | | |
| Emergency department visit for infection – last 12 months | | | | | |
| Hospitalisation for infection – first 5 years of life | | | | | |
| Hospitalisation for infection – last 12 months | | | | | |

### Slide 11
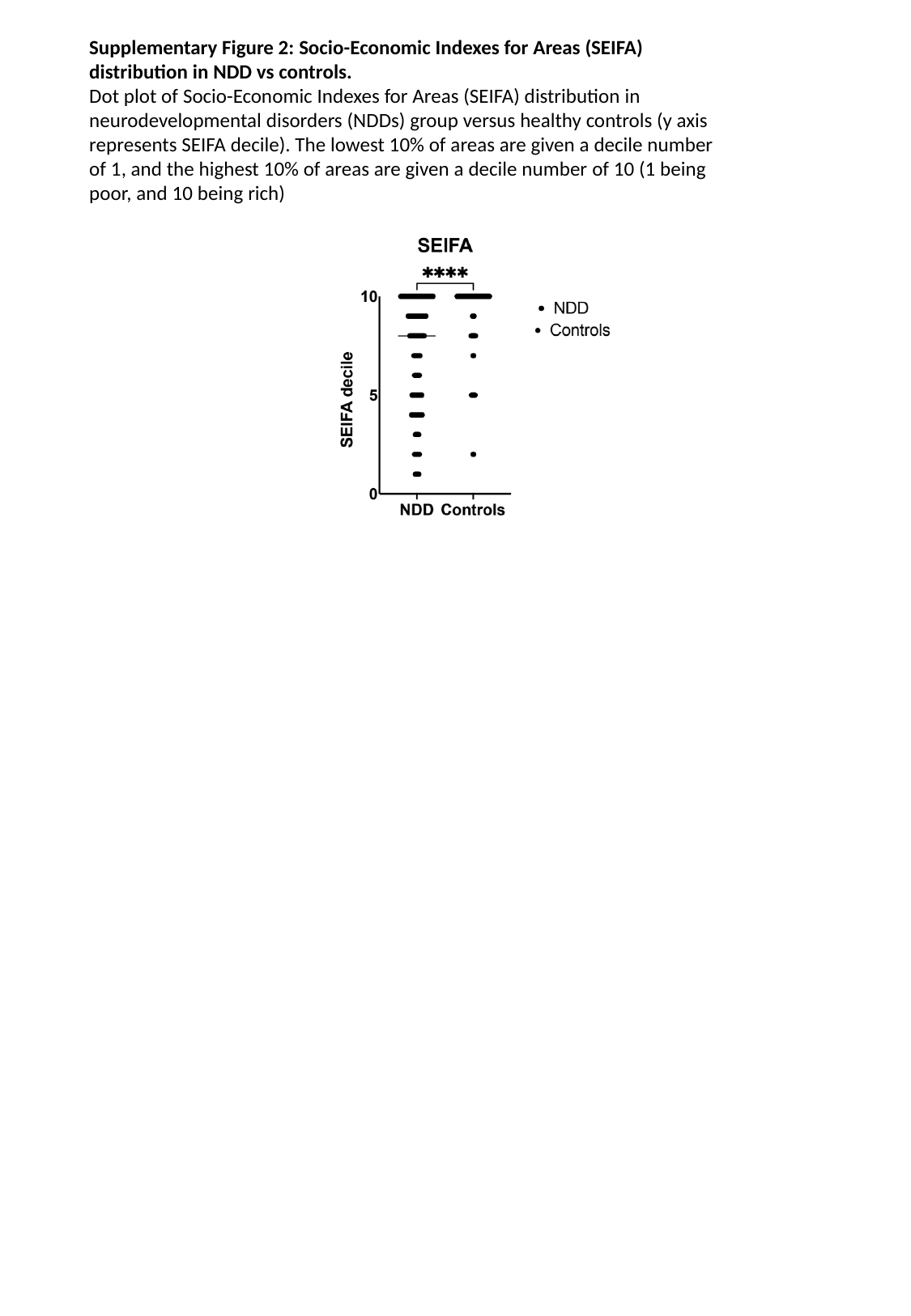

Supplementary Figure 2: Socio-Economic Indexes for Areas (SEIFA) distribution in NDD vs controls.
Dot plot of Socio-Economic Indexes for Areas (SEIFA) distribution in neurodevelopmental disorders (NDDs) group versus healthy controls (y axis represents SEIFA decile). The lowest 10% of areas are given a decile number of 1, and the highest 10% of areas are given a decile number of 10 (1 being poor, and 10 being rich)

### Slide 12
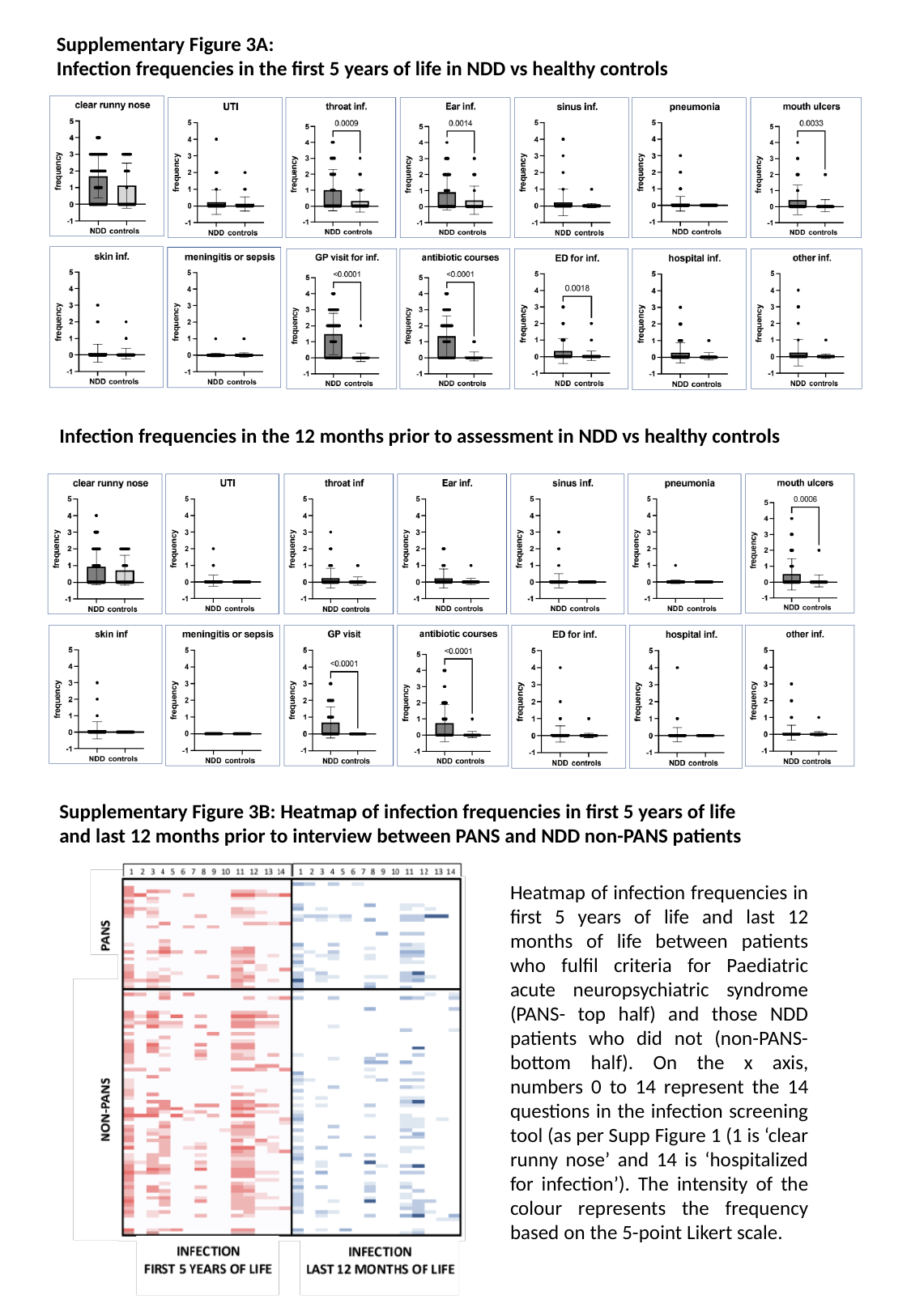

Supplementary Figure 3A:
Infection frequencies in the first 5 years of life in NDD vs healthy controls
Infection frequencies in the 12 months prior to assessment in NDD vs healthy controls
Supplementary Figure 3B: Heatmap of infection frequencies in first 5 years of life and last 12 months prior to interview between PANS and NDD non-PANS patients
Heatmap of infection frequencies in first 5 years of life and last 12 months of life between patients who fulfil criteria for Paediatric acute neuropsychiatric syndrome (PANS- top half) and those NDD patients who did not (non-PANS- bottom half). On the x axis, numbers 0 to 14 represent the 14 questions in the infection screening tool (as per Supp Figure 1 (1 is ‘clear runny nose’ and 14 is ‘hospitalized for infection’). The intensity of the colour represents the frequency based on the 5-point Likert scale.

### Slide 13
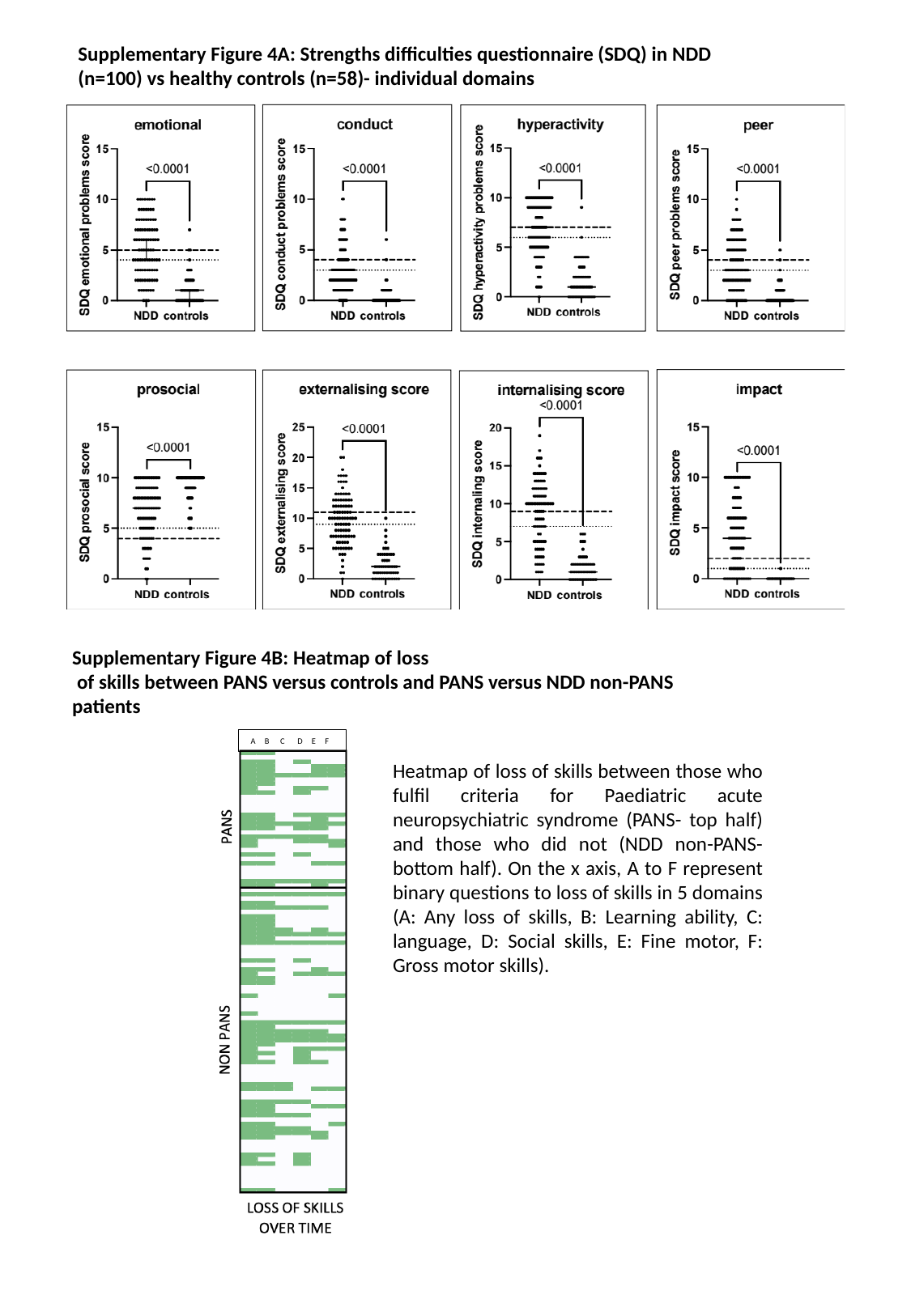

Supplementary Figure 4A: Strengths difficulties questionnaire (SDQ) in NDD (n=100) vs healthy controls (n=58)- individual domains
Supplementary Figure 4B: Heatmap of loss
 of skills between PANS versus controls and PANS versus NDD non-PANS patients
A B C D E F
Heatmap of loss of skills between those who fulfil criteria for Paediatric acute neuropsychiatric syndrome (PANS- top half) and those who did not (NDD non-PANS- bottom half). On the x axis, A to F represent binary questions to loss of skills in 5 domains (A: Any loss of skills, B: Learning ability, C: language, D: Social skills, E: Fine motor, F: Gross motor skills).

### Slide 14
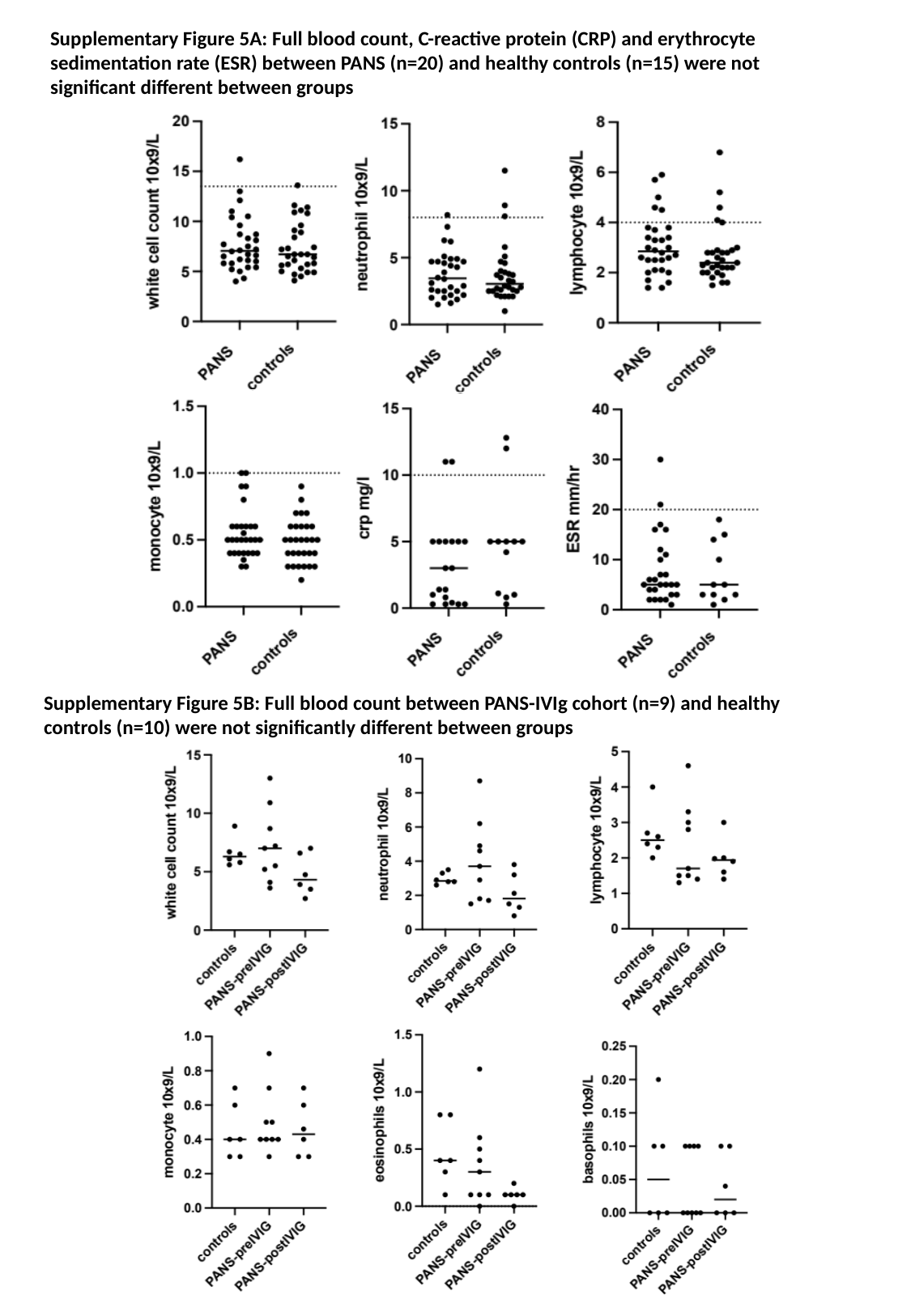

Supplementary Figure 5A: Full blood count, C-reactive protein (CRP) and erythrocyte sedimentation rate (ESR) between PANS (n=20) and healthy controls (n=15) were not significant different between groups
Supplementary Figure 5B: Full blood count between PANS-IVIg cohort (n=9) and healthy controls (n=10) were not significantly different between groups

### Slide 15
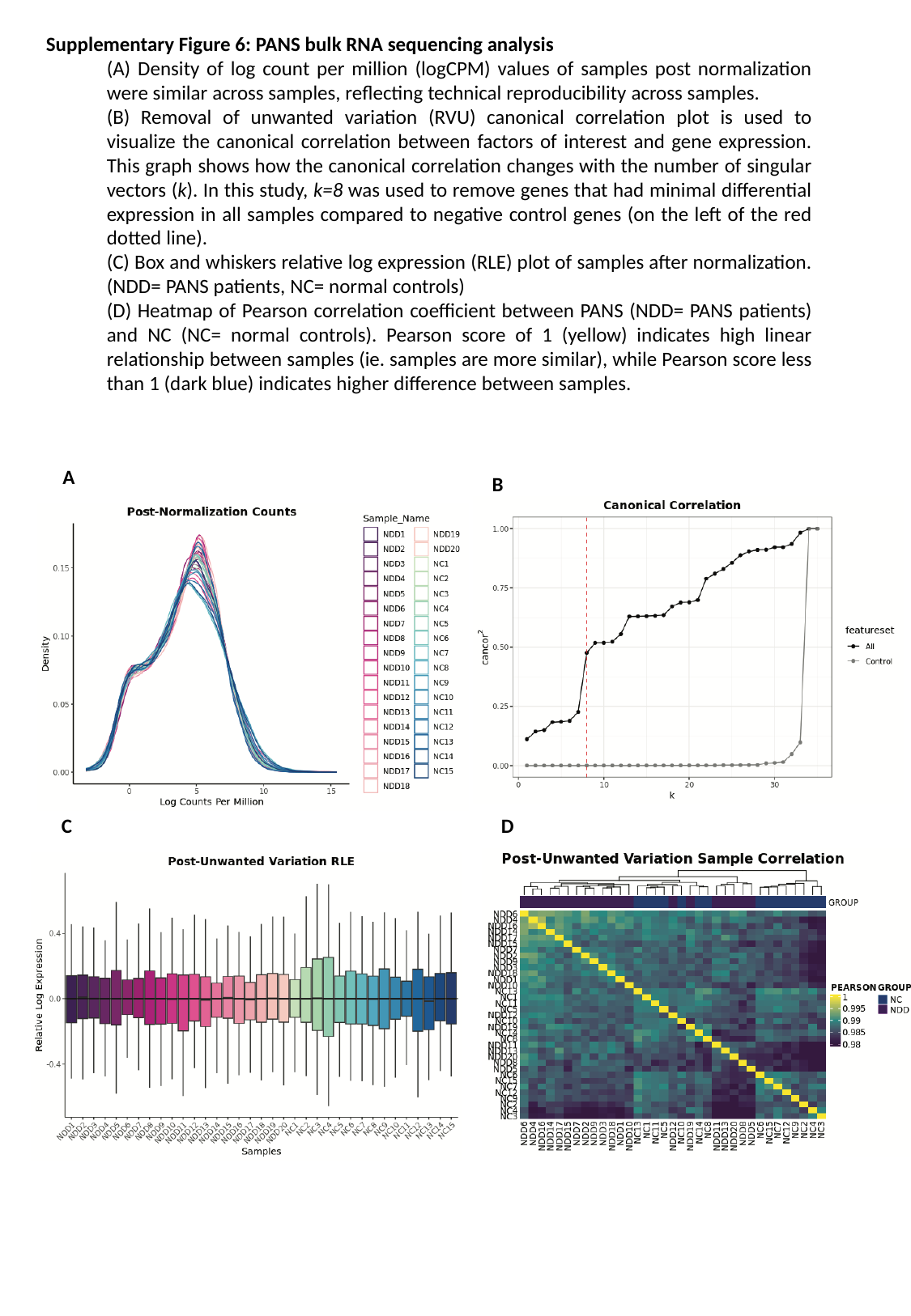

Supplementary Figure 6: PANS bulk RNA sequencing analysis
(A) Density of log count per million (logCPM) values of samples post normalization were similar across samples, reflecting technical reproducibility across samples.
(B) Removal of unwanted variation (RVU) canonical correlation plot is used to visualize the canonical correlation between factors of interest and gene expression. This graph shows how the canonical correlation changes with the number of singular vectors (k). In this study, k=8 was used to remove genes that had minimal differential expression in all samples compared to negative control genes (on the left of the red dotted line).
(C) Box and whiskers relative log expression (RLE) plot of samples after normalization. (NDD= PANS patients, NC= normal controls)
(D) Heatmap of Pearson correlation coefficient between PANS (NDD= PANS patients) and NC (NC= normal controls). Pearson score of 1 (yellow) indicates high linear relationship between samples (ie. samples are more similar), while Pearson score less than 1 (dark blue) indicates higher difference between samples.
A
B
C
D

### Slide 16
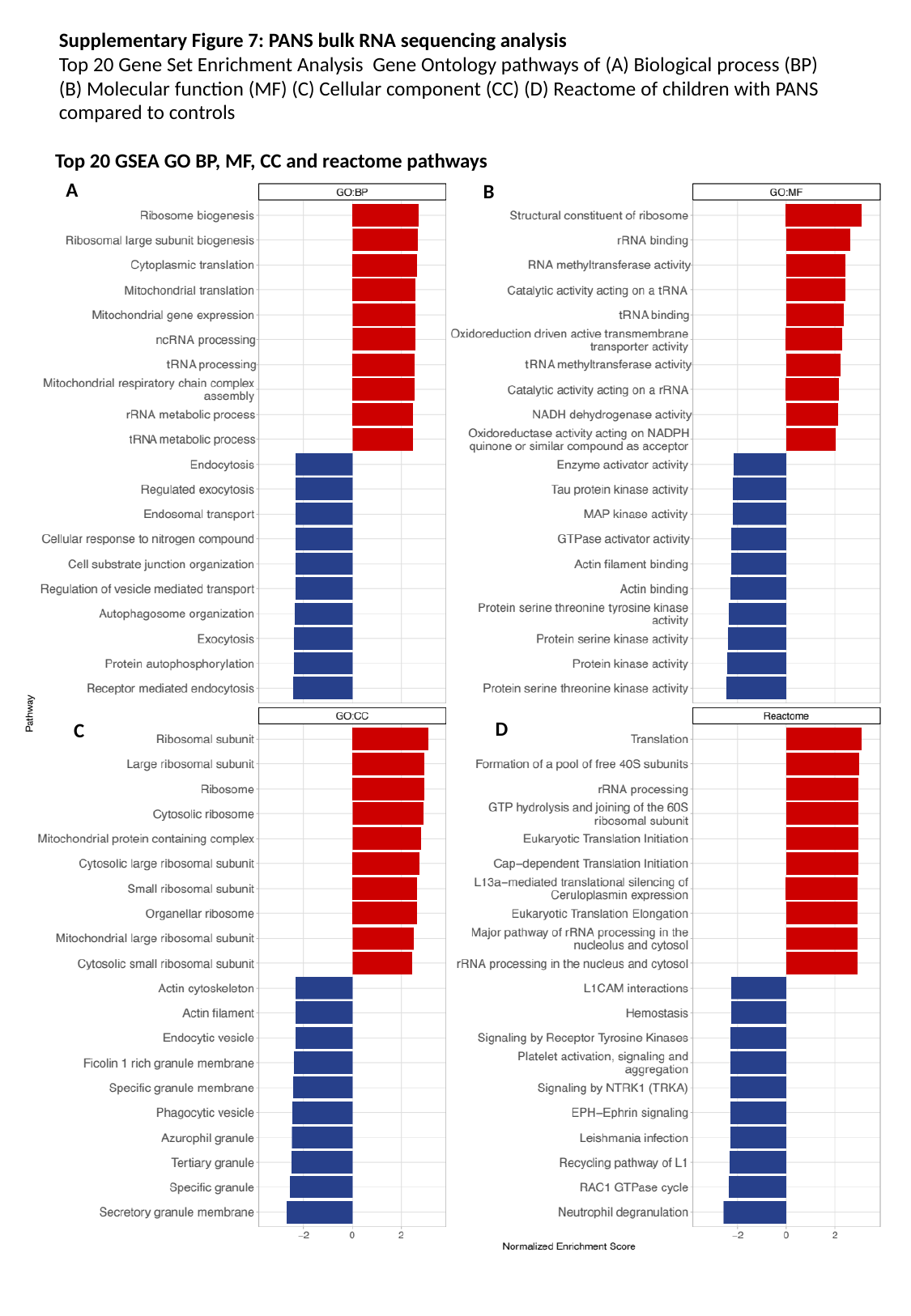

Supplementary Figure 7: PANS bulk RNA sequencing analysis
Top 20 Gene Set Enrichment Analysis Gene Ontology pathways of (A) Biological process (BP) (B) Molecular function (MF) (C) Cellular component (CC) (D) Reactome of children with PANS compared to controls
Top 20 GSEA GO BP, MF, CC and reactome pathways
A
B
D
C

### Slide 17
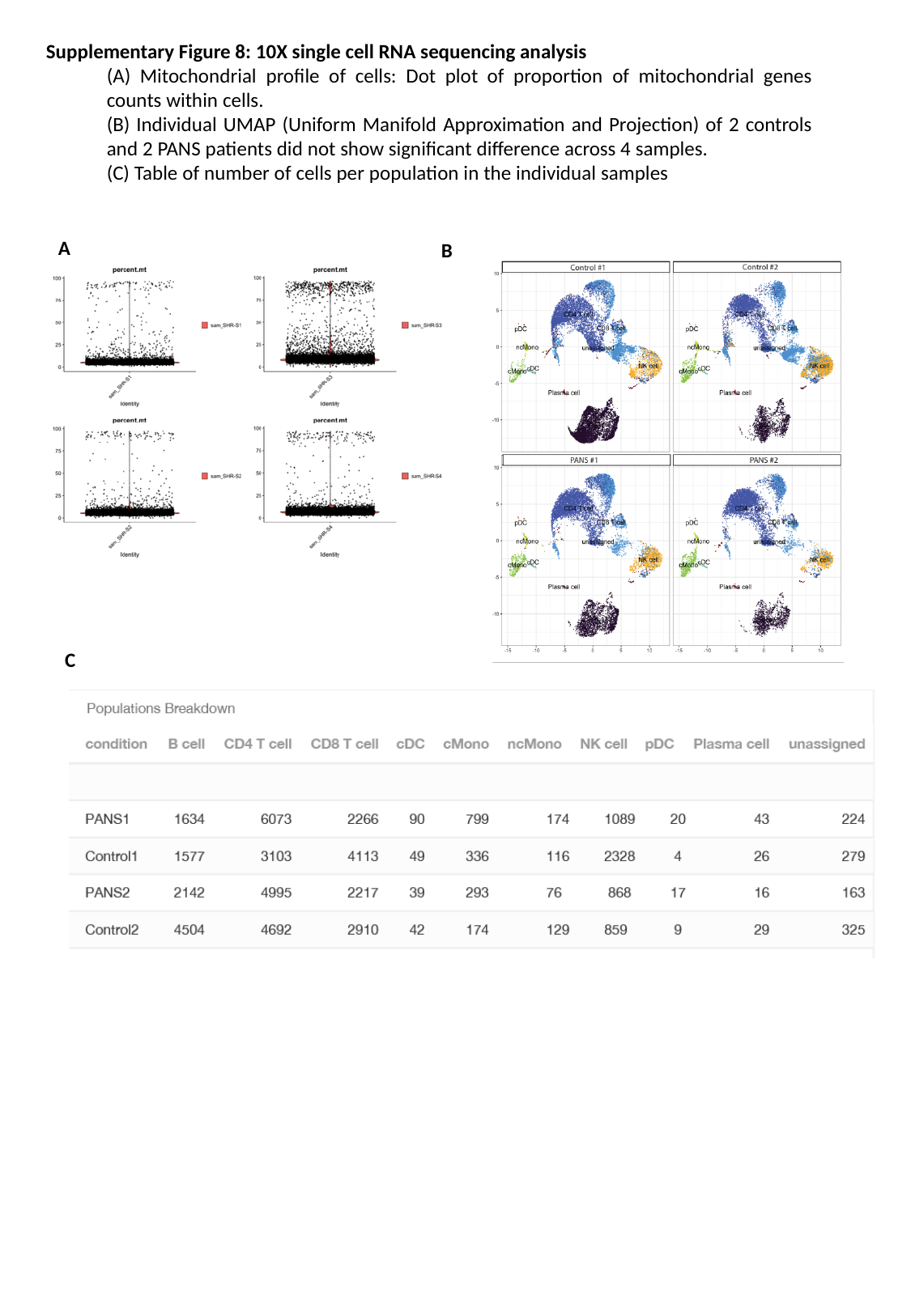

Supplementary Figure 8: 10X single cell RNA sequencing analysis
(A) Mitochondrial profile of cells: Dot plot of proportion of mitochondrial genes counts within cells.
(B) Individual UMAP (Uniform Manifold Approximation and Projection) of 2 controls and 2 PANS patients did not show significant difference across 4 samples.
(C) Table of number of cells per population in the individual samples
A
B
C

### Slide 18
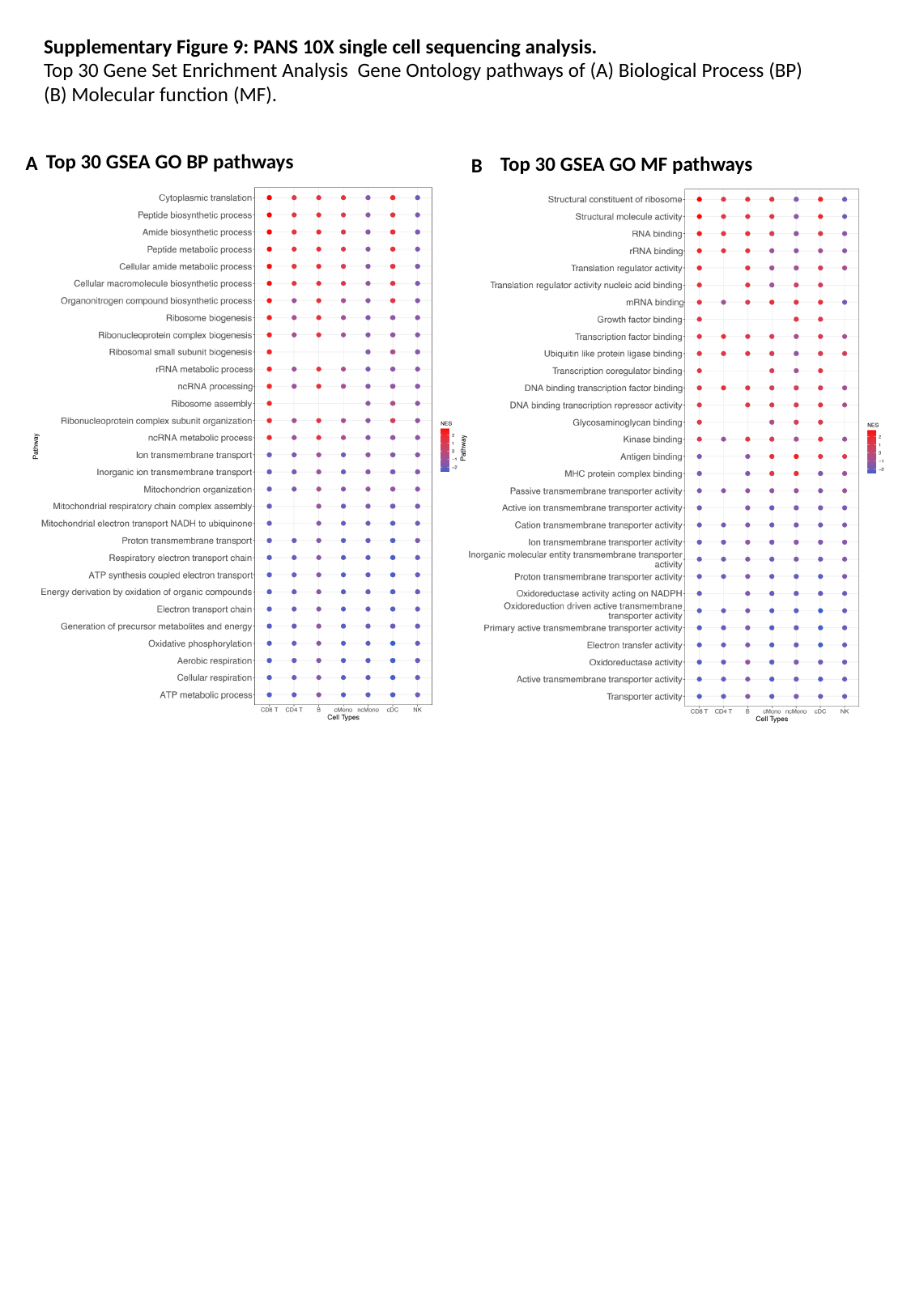

Supplementary Figure 9: PANS 10X single cell sequencing analysis.
Top 30 Gene Set Enrichment Analysis Gene Ontology pathways of (A) Biological Process (BP) (B) Molecular function (MF).
Top 30 GSEA GO BP pathways
A
Top 30 GSEA GO MF pathways
B

### Slide 19
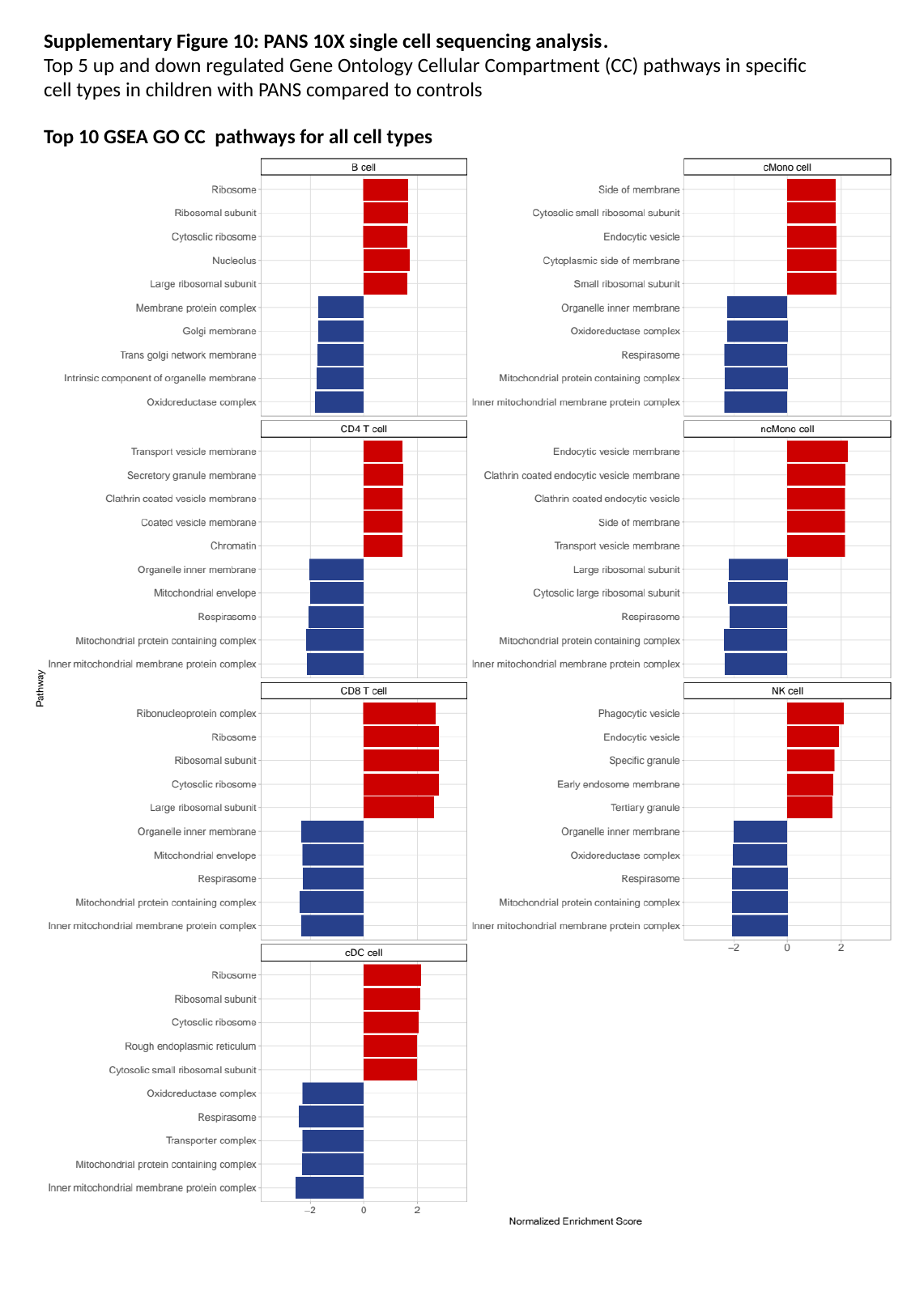

Supplementary Figure 10: PANS 10X single cell sequencing analysis.
Top 5 up and down regulated Gene Ontology Cellular Compartment (CC) pathways in specific cell types in children with PANS compared to controls
Top 10 GSEA GO CC pathways for all cell types

### Slide 20
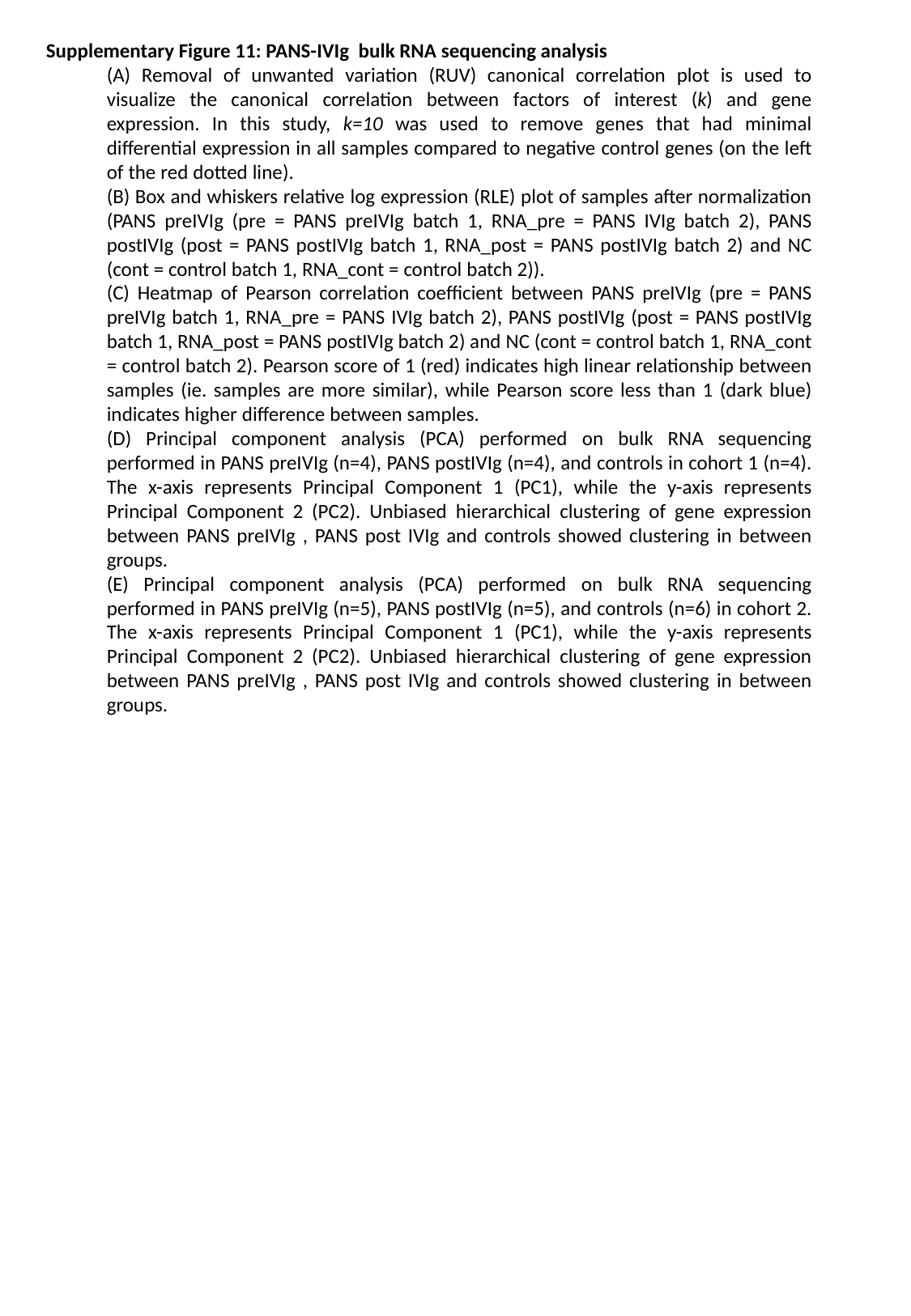

Supplementary Figure 11: PANS-IVIg bulk RNA sequencing analysis
(A) Removal of unwanted variation (RUV) canonical correlation plot is used to visualize the canonical correlation between factors of interest (k) and gene expression. In this study, k=10 was used to remove genes that had minimal differential expression in all samples compared to negative control genes (on the left of the red dotted line).
(B) Box and whiskers relative log expression (RLE) plot of samples after normalization (PANS preIVIg (pre = PANS preIVIg batch 1, RNA_pre = PANS IVIg batch 2), PANS postIVIg (post = PANS postIVIg batch 1, RNA_post = PANS postIVIg batch 2) and NC (cont = control batch 1, RNA_cont = control batch 2)).
(C) Heatmap of Pearson correlation coefficient between PANS preIVIg (pre = PANS preIVIg batch 1, RNA_pre = PANS IVIg batch 2), PANS postIVIg (post = PANS postIVIg batch 1, RNA_post = PANS postIVIg batch 2) and NC (cont = control batch 1, RNA_cont = control batch 2). Pearson score of 1 (red) indicates high linear relationship between samples (ie. samples are more similar), while Pearson score less than 1 (dark blue) indicates higher difference between samples.
(D) Principal component analysis (PCA) performed on bulk RNA sequencing performed in PANS preIVIg (n=4), PANS postIVIg (n=4), and controls in cohort 1 (n=4). The x-axis represents Principal Component 1 (PC1), while the y-axis represents Principal Component 2 (PC2). Unbiased hierarchical clustering of gene expression between PANS preIVIg , PANS post IVIg and controls showed clustering in between groups.
(E) Principal component analysis (PCA) performed on bulk RNA sequencing performed in PANS preIVIg (n=5), PANS postIVIg (n=5), and controls (n=6) in cohort 2. The x-axis represents Principal Component 1 (PC1), while the y-axis represents Principal Component 2 (PC2). Unbiased hierarchical clustering of gene expression between PANS preIVIg , PANS post IVIg and controls showed clustering in between groups.

### Slide 21
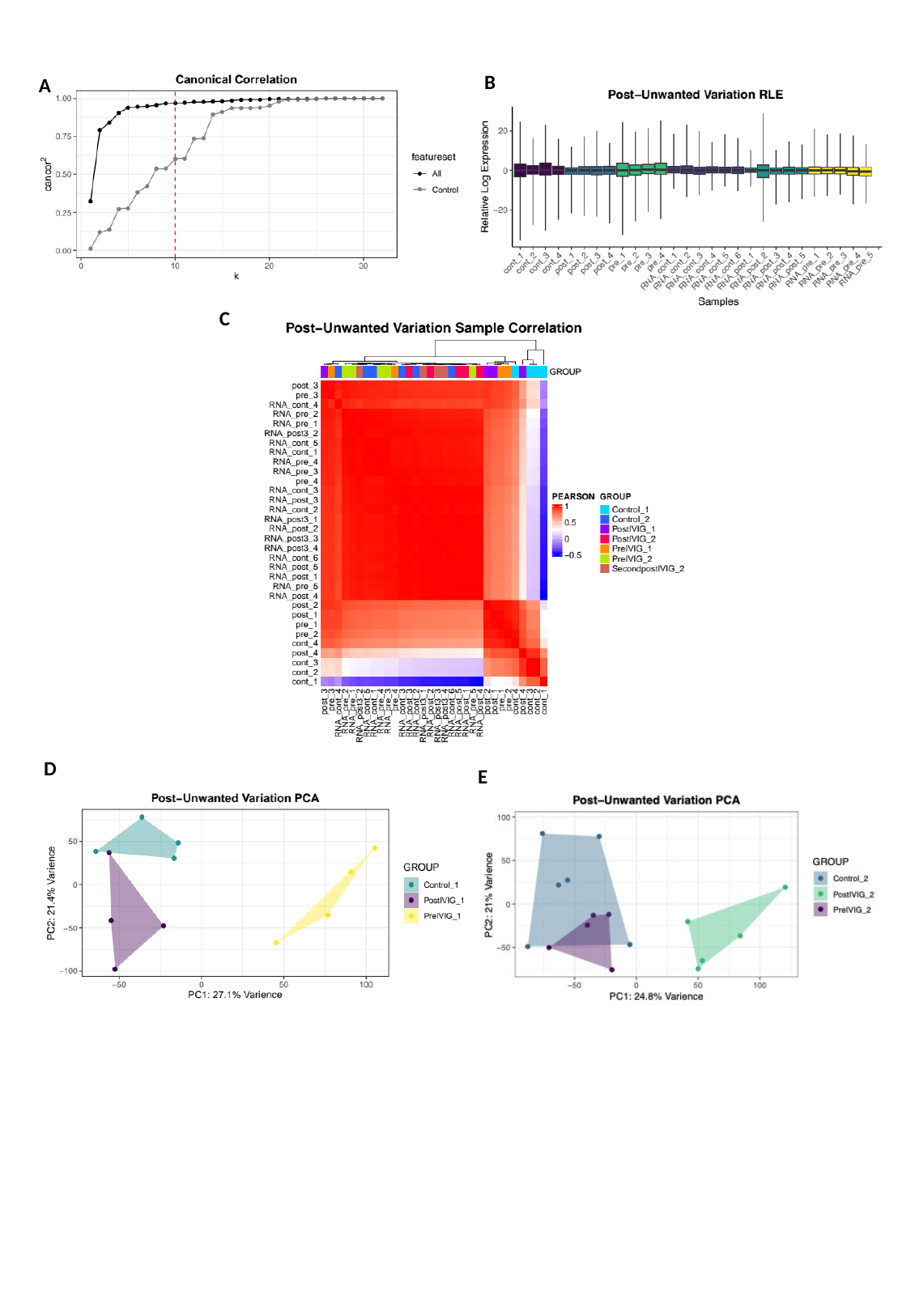

B
A
C
D
E

### Slide 22
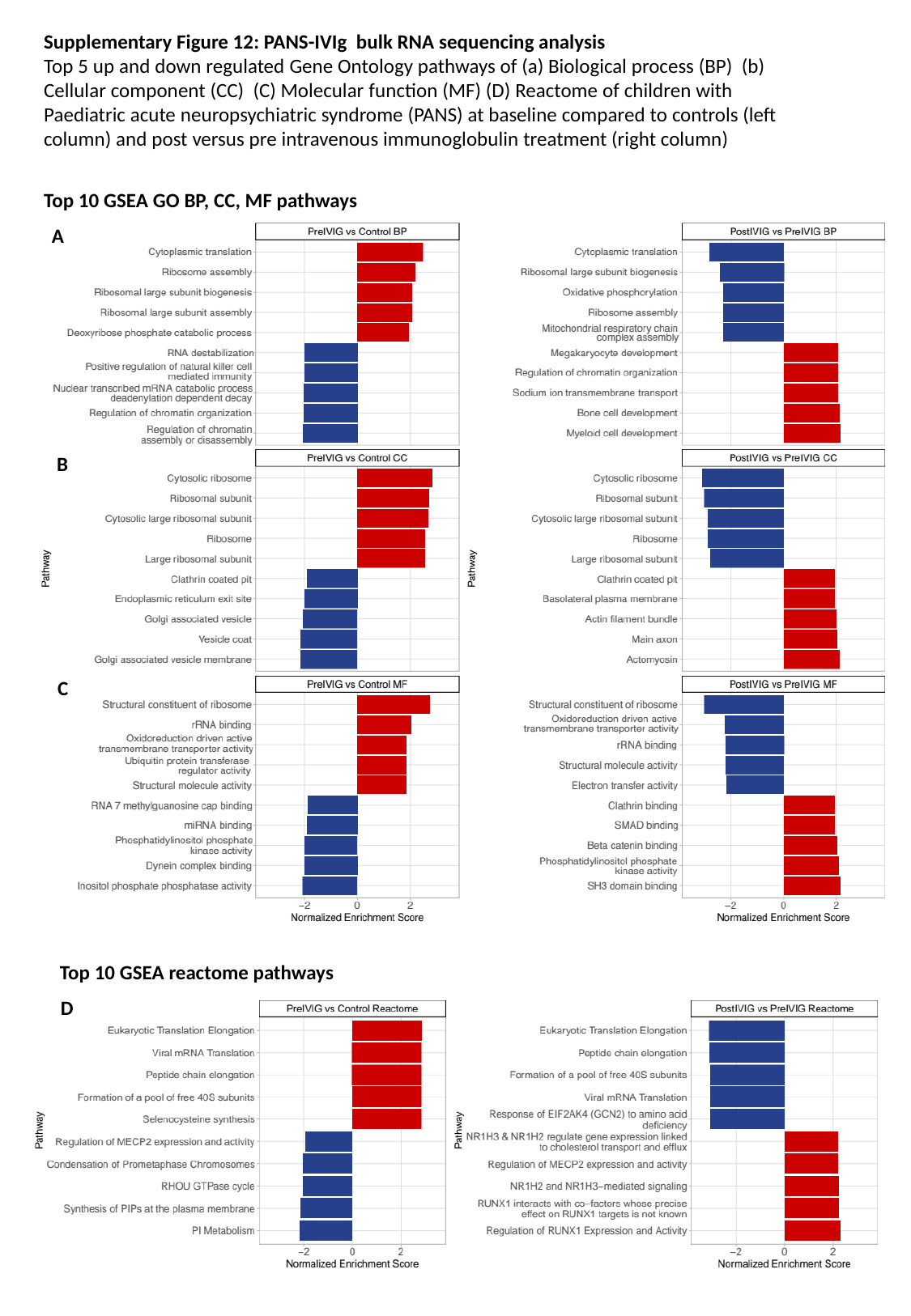

Supplementary Figure 12: PANS-IVIg bulk RNA sequencing analysis
Top 5 up and down regulated Gene Ontology pathways of (a) Biological process (BP) (b) Cellular component (CC) (C) Molecular function (MF) (D) Reactome of children with Paediatric acute neuropsychiatric syndrome (PANS) at baseline compared to controls (left column) and post versus pre intravenous immunoglobulin treatment (right column)
Top 10 GSEA GO BP, CC, MF pathways
A
B
C
Top 10 GSEA reactome pathways
D
